# 2.5 Million Person-Years of Life Have Been Lost Due to COVID-19 in the United States

**DOI:** 10.1101/2020.10.18.20214783

**Authors:** Stephen J. Elledge

## Abstract

The COVID-19 pandemic, caused by tens of millions of SARS-CoV-2 infections world-wide, has resulted in considerable levels of mortality and morbidity. The United States has been hit particularly hard having 20 percent of the world’s infections but only 4 percent of the world population. Unfortunately, significant levels of misunderstanding exist about the severity of the disease and its lethality. As COVID-19 disproportionally impacts elderly populations, the false impression that the impact on society of these deaths is minimal may be conveyed by some because elderly individuals are closer to a natural death. To assess the impact of COVID-19 in the US, I have performed calculations of person-years of life lost as a result of 194,000 premature deaths due to SARS-CoV-2 infection as of early October, 2020. By combining actuarial data on life expectancy and the distribution of COVID-19 associated deaths we estimate that over 2,500,000 person-years of life have been lost so far in the pandemic in the US alone, averaging over 13.25 years per person with differences noted between males and females. Importantly, nearly half of the potential years of life lost occur in non-elderly populations. Issues impacting refinement of these models and the additional morbidity caused by COVID-19 beyond lethality are discussed.

## Introduction

Coronavirus Disease 19 (COVID-19), the disease resulting from SARS-CoV-2 infection, is the greatest global health crisis since the 1918 influenza pandemic and has caused significant mortality and morbidity throughout the world (Hu et al., 2020) as well as severe economic disruption. The COVID-19 pandemic started in Wuhan, China, with the first reported case on Dec. 8, 2019 then rapidly spread throughout Europe. Since then nearly 40 million cases have been documented world-wide with over 1 million deaths, affecting individual countries to varying degrees. For example, Italy and Spain were profoundly affected while countries like South Korea experienced substantially less morbidity and mortality (Issac et al., 2020). The United States was hit especially hard, experiencing over 20% of the world’s infections and deaths while possessing only 4% of the world’s population (JHU [Johns Hopkins University] Coronavirus Resource Center, https://coronavirus.jhu.edu/map.html).

While differences in population structures, demographics, genetics and mitigation efforts are likely to explain the distinct outcomes experienced across different countries, some aspects of COVID-19 are shared across countries. These commonalities include severity and lethality with respect to sex, age and ethnicity. Ethnicity is complicated by socioeconomic status and population density of living circumstances, which is well known to impact the incidence of viral infection, for example CMV (Lachmann et al., 2018; Bate et al., 2010). COVID-19 causes significant mortality and morbidity. The clinical course of COVID-19 is notable for its extreme variability: while some individuals remain entirely asymptomatic, others experience fever, anosmia, diarrhea, severe respiratory distress, pneumonia, cardiac arrhythmia, blood clotting disorders, liver and kidney distress, enhanced cytokine release and death (Yuki et al., 2020). Some of these infections result in long-lasting disabilities (Cousin-Frankel, 2020), the magnitude of which will only become clear over a longer time horizon.

Quantifying the impact of the COVID-19 pandemic is critical for the public and policy makers to be properly informed as to the societal cost of the pandemic in order to rationally determine how best to minimize the social costs of the disease. Significant levels of misunderstanding exist about the severity of the disease and its lethality. For example, because the great majority of COVID-19 deaths occur among the elderly (Wortham et al., 2020), the false impression that the impact on society from these deaths is minimal may be conveyed since these individuals were closer to a natural death. Aside from any troubling ethical implications associated with rationalization of COVID-19 mortality along these lines, such a conclusion is unwarranted for at least two reasons. First, as individuals age, their life expectancies increase too, well beyond the life expectancy at birth, which is the value most familiar to the general public. Second, a significant number of relatively young individuals have also died from COVID-19 and had decades of remaining life expectancy. Case fatality rates and total mortality are inadequate measures to portray the true impact of the disease on a population. In order to provide a better metric for the demographic impact of COVID-19 in the United States, I calculate the Potential Years of Life Lost (PYLL) for COVID-19 in the US using the most recent death counts stratified for sex and age. These analyses show that rather than minimal effects on society, over 2.5 million years of life have been forfeited due to the COVID-19 epidemic in the United States.

## Results

To calculate the person-years of life lost in the US, I first obtained the publicly available actuarial tables from the Social Security Administration web site (Actuarial Life Table, https://www.ssa.gov/oact/STATS/table4c6.html#fn1) to examine the normal life expectancies in the US across different age groups by sex (Table 1). I next consulted the publicly available Centers for Disease Control and Prevention (CDC) reports of deaths per age group present on the CDC website on Oct. 3, 2020 (Table 2) (COVID-19 Deaths in the US, CDC as of OCT. 3 2020 https://www.cdc.gov/nchs/nvss/vsrr/covid_weekly/index.htm). Data on COVID-19-related deaths is presented in 10-year spans in the CDC table. I calculated the average life expectancy for each span of 10 years presented. Because of the differences in mortality and morbidity between males and females, the genders were analyzed separately. This was accomplished by analysis of the distribution of life expectancies of the given span from Table 2, summing them within an indicated span of years and taking the average for that group of years (Table 3). For example, the life expectancies calculated as expected remaining years for the age group 45-54 in females ranges from 37.80-29.68 additional years with an average of 33.704 years. In order to make a simple calculation of person-years lost to COVID-19, the average life expectancy per age group was multiplied by the number of deaths in that age group. The person-years of lost life was then summed across all ages to determine the total person-years lost (Table 4). For the 104,896 deaths in males, the total person-years lost was 1,461,662. For the 89,191 deaths in females, the total person-years lost was 1,110,440. Altogether, 2,572,102 person-years were lost due to a total of 194,087 deaths. The average number of person-years lost per individual death was 13.25, with males losing 13.93 years of life per death and females losing 12.45 years of life.

**Figure 1.**
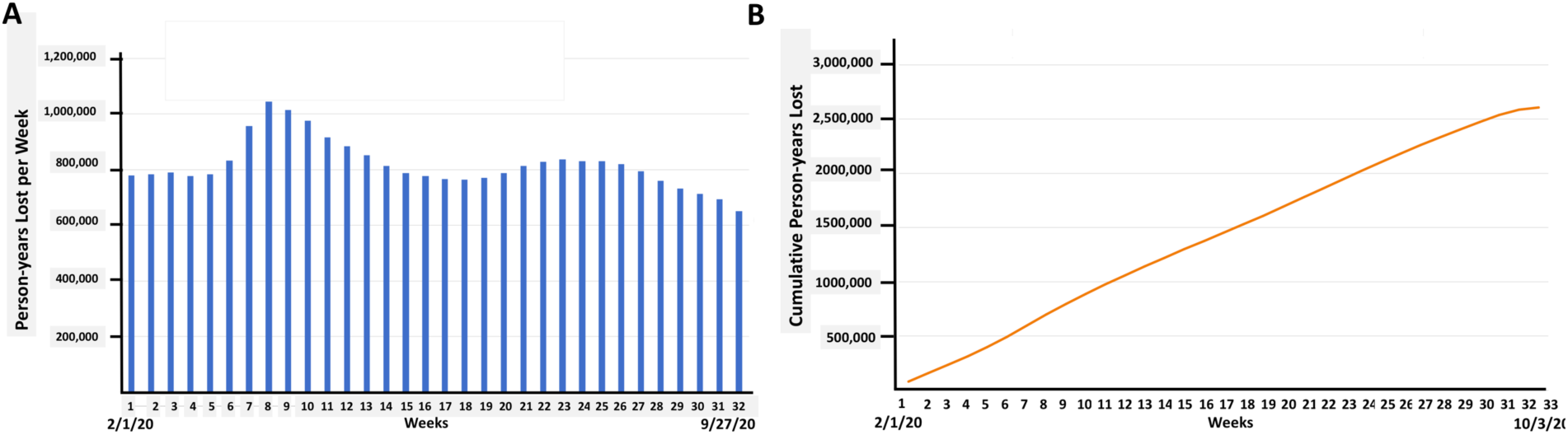
Person-years lost per week of the pandemic due to COVID-19 associated deaths. (A) Person-years lost per week through 9/27/20. (B) Cumulative person-years lost through 10/3/20. Weekly totals were from https://www.cdc.gov/nchs/nvss/vsrr/COVID19/index.htm

### Potential sources of error

#### Age distributions

Age is a prominent comorbidity factor in the death rate from COVID-19. It may be unrealistic to assume that there is an even distribution of deaths across specific 10-year spans that form the CDC groups. More realistically, the probability of death due to COVID-19 infection, beyond some benchmark age, increases slightly every year. This means that it may be that more deaths are distributed at the higher end of the age range within a decade than at the beginning where younger individuals reside, a fact not yet fully accounted for in this analysis. To begin to arrive at an understanding of how death rates are distributed according to age, I normalized the number of deaths per age group to the number of living individuals in the US in that age range by calculating the deaths per million in that group (Table 5A). Although I did not have information on the distribution of deaths within age groups, with the rates normalized to population as deaths per million, I could determine how death rates increased from one age group to the next and whether there was a pattern that would facilitate further analysis. Surprisingly, the increased probability of dying in the next decade was remarkably stable from one decade to the next. Deaths among very young COVID-19-infected individuals are anomalies and cannot be reliably factored into the overall calculations as they account for such a small fraction of total deaths. For example, deaths among females younger than age 35 make up only 0.75% of the total COVID-19 deaths. However, the increased probability from one decade of individuals to the next after age 35 was largely uniform within a given gender and averaged 2.4-fold for men and 2.7-fold for women (Table 5A). Since the fold increase in probability of death was uniform from one decade to the next it was also likely to be relatively uniform from one year to the next. This observation then allowed the distribution of deaths within a decade from year to year to better model the loss of life expectancy calculated previously (Table 6). This averaged out to a 1.104-fold increase per year (or a 10.4% rise per year) for men and a 1.091-fold increase per year (or a 9.14% rise per year) for women (Table 5C). This number was used to distribute the deaths per age range to year-by-year deaths. This was accomplished similarly to compound interest whereby a base number of deaths for year was multiplied by 1 for the first year, then for males 1.104 for year two, then that number was multiplied by a factor of 1.104 again, and so on until the tenth year. Mathematically, the expression for these calculations is given by:

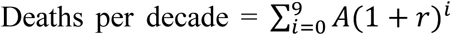, where A is the base number of deaths, *r* is the percentage annual increase in deaths imputed from the decadal rate and represented by the decimal, i.e. 10% = 0.10, and *i* the year with 0 as a base.

To arrive at the adjusted age distribution of person-years lost for each year, three numbers are multiplied; the base number, the appropriate factor, and the remaining life expectancy for that year. The exception to this formula is the 1-4 years old group in which we simply divided the number of deaths by 4 and used the same number for each year since there was no reason to suspect a gradation in deaths at that young age. Since the CDC group deaths for those ages 85 and above as a single number, we took that number and included all deaths in the 85-94 category and generated a base number and factor covering all deaths into that 10-year period. The fraction of the total population in the US that is 95 or older is approximately 7.7% of those 85 or older according to the 2010 US Census (Table 7). Thus, of the 60,000 deaths in this total age group that have occurred due to COVID-19, 4,200 deaths would be estimated to occur in the 95 or older age group if death rates were approximately equal between the two groups. If we estimate they are 2.5-fold greater in the latter that would be approximately 10,000 deaths whose person years are over estimated by their inclusion in the 85-94 group, perhaps by 2 years or a total of 20,000 person-years which is a minor error on the total of 2.5 million person-years (see discussion in Table 4).

Performing the age distribution adjustment as described above is shown in Table 6. The number of person-years lost is reduced by 3.0% for males and 3.7% for females, resulting in total person-years lost of 2,486,160 rather than 2,572,102.

#### Ethnicity distributions among fatalities

A second source of potential error concerns the different life expectancies experienced by the ethnic groups that make up the US population. According to the CDC, deaths due to COVID-19 occur more frequently in Black and Latinx populations relative to non-Hispanic Whites. This is separate from the potential enhanced mortality that occurs in minority populations (Alcendor et al., 2020, Phillips et al., 2020). Current estimates are that Whites correspond to approximately 51% of deaths with 20% in Blacks, 21% in Hispanic or Latinx populations and approximately 8% in a group labelled “other” that include Asians, Polynesian and unidentified (Table 8). Since we do not have age groupings for these deaths based on ethnic identity for the same date of October 3, 2020 and we do not have actuarial tables for all categories of ethnicity, it is not possible to perform precisely the same analysis as shown above in Table 6 for each individual ethnic group to determine potential years of life lost. However, this is unlikely to present a major problem with the analysis above because when one looks at the differences in the life expectancies of the dominant groups in the study that account for >90% of the deaths, Non-Hispanic Whites, Non-Hispanic Blacks, and Hispanic populations, a pattern emerges in that the life expectancies for Blacks are lower than Whites by about the same degree as the life expectancy for Hispanics is above Whites. This is true for both males and females. Therefore, since the number of deaths is approximately equal between Black and Hispanics and their death distribution according to ages are similar, their effects cancel each other out and average out to be approximately what the Non-Hispanic White life expectancy is (Table 9). In fact, the average of the non-Hispanic blacks and Hispanics adds half of a year to life expectancy for a given age, which would increase the person-years lost on average. In addition, there is no information on the life expectancy of the heterogeneous group labeled “Other” which is small comprising only 8% of the deaths. How that impacts the calculation of person-years lost is not known but is likely to be minor.

## Discussion

The total number of deaths attributed to the COVID-19 pandemic is growing at an alarming rate. In this study I have attempted to quantify the extent of life lost so far in this pandemic as of October 3, 2020 where data for the age and sex distribution of over 194,000 COVID-19 associated deaths was available. These calculations reveal a profound loss of life as measured in person-years of almost 2.5 million person-years as of early October, 2020 in the United States. This corresponds to an average loss of life of 13.25 person-years per COVID-19 associated death. This is an astounding cost and surprising given the apparent public misperception that COVID-19 is a disease that disproportionately impacts the elderly and is somehow of less concern to the rest of society. This misunderstanding is a result of a failure to appreciate the fact that individuals considered elderly still have substantial remaining life expectancies relative to the life expectancy at birth. Secondly, a significant proportion of deaths due to COVID-19 occur in individuals in their 40s, 50s and 60s who had dozens of years of expected life ahead of them.

Several previous studies have been performed on this topic and arrived at similar conclusions with respect to person-years lost proportional to the number of deaths at the time of their analysis (Mitra et al., 2020; Oh et al., 2020; Goldstein et al., 2020). They project averages with slightly fewer person-years lost per death. It is not known if those subtle differences with this study have to do with the methods of analysis which appear to be similar in principle but distinct in details of the assumptions employed in the models, or to the fact that several of the previous studies were performed earlier in the pandemic when large segments of the lives lost were from elderly care facilities and thus the average ages were older. If the mortality has shifted to a proportionally younger demographic due to increased incidence, additional deaths will render a greater cost in person years of life lost. Already in this analysis 45% of the person-years lost are between the ages of 0 and 64 overall, and 50% of the person-years lost in men fall in that age range.

**Figure 2.**
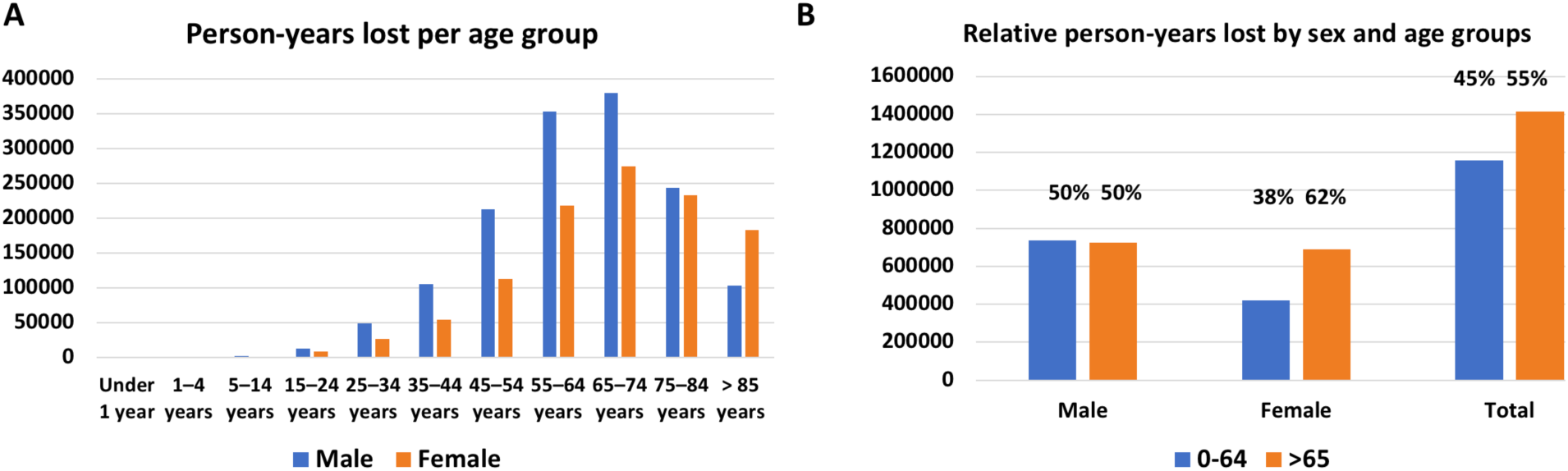
PYLL distribution by sex and age. (**A**) Person-yeast lost by age. (**B**) PYLL by sex.

An important variable that this and other studies have not been able to adequately incorporate into this analysis is the effect of comorbidities on life expectancy of COVID-19 deaths which is due to a lack of appropriate statistical information. This must, therefore, be considered a potential source of error. The SARS-CoV-2 virus is known to infect and replicate in many different tissues and exacerbates problems in several organ systems including the kidney, liver, heart, lungs and brain (Lu et al., 2020; Chandrashekar et al., 2020). Any individual with problems in these systems or the immune system is likely to be more vulnerable to SARS-CoV-2 infection and suffer more severe outcomes as has been demonstrated for immune deficiencies (Bastard et al., 2020). In addition, other health states qualifying as pre-existing conditions, such as obesity, hypertension, chronic kidney disease and diabetes are known comorbidity factors for COVID-19 (see CDC co-morbidity tables and references therein; https://www.cdc.gov/coronavirus/2019-ncov/need-extra-precautions/evidence-table.html) and these cohorts of individuals have a shorter than average predicted life span. Deaths due to complications with pre-existing comorbid conditions would artificially increase the person-years lost in these calculations but are difficult to quantitate in this current analysis. However, most people over the age of 60 have comorbidities so these are already factored into the longevity tables. For example, one of the largest co-morbidity factors is diabetes (Table 10, 33,100 deaths out of 201,000, or 16.5% of COVID-19 deaths) and diabetes is thought to shorten life expectancy by approximately 8 years per individual and has lowered overall average life expectancy in the US by 0.83 and 0.89 years for males and females, respectively (Preston et al., 2018b). However, the prevalence of diabetes is 10.4% in the general population and its effects on life expectancy has already been factored into the actuarial tables. Thus, approximately 6% of the 194,000 COVID-19 deaths in our analysis, 11,640 in total, are likely due to excess diabetes deaths over the average amount in the population. This would result in an excess of 93,120 person-years, but this would need to be further adjusted for excess deaths per age because on average people over 80 years old have less than 8 years of residual life expectancy. As a result, diabetes may have less than a 4% impact on the current estimates as not all deaths are in individuals with a known co-morbidity. However, other comorbidities listed in

Table 10 are actually at a lower prevalence than in the general population, i.e. Alzheimer’s disease and hypertension, and could be due to undercounting due to multiple comorbidities. Thus, it is not yet clear how to fully interpret these data and adjust our calculations. We include a 15% estimate of reduction in PYLL in the summary table (Table 11).

**Table 11.**
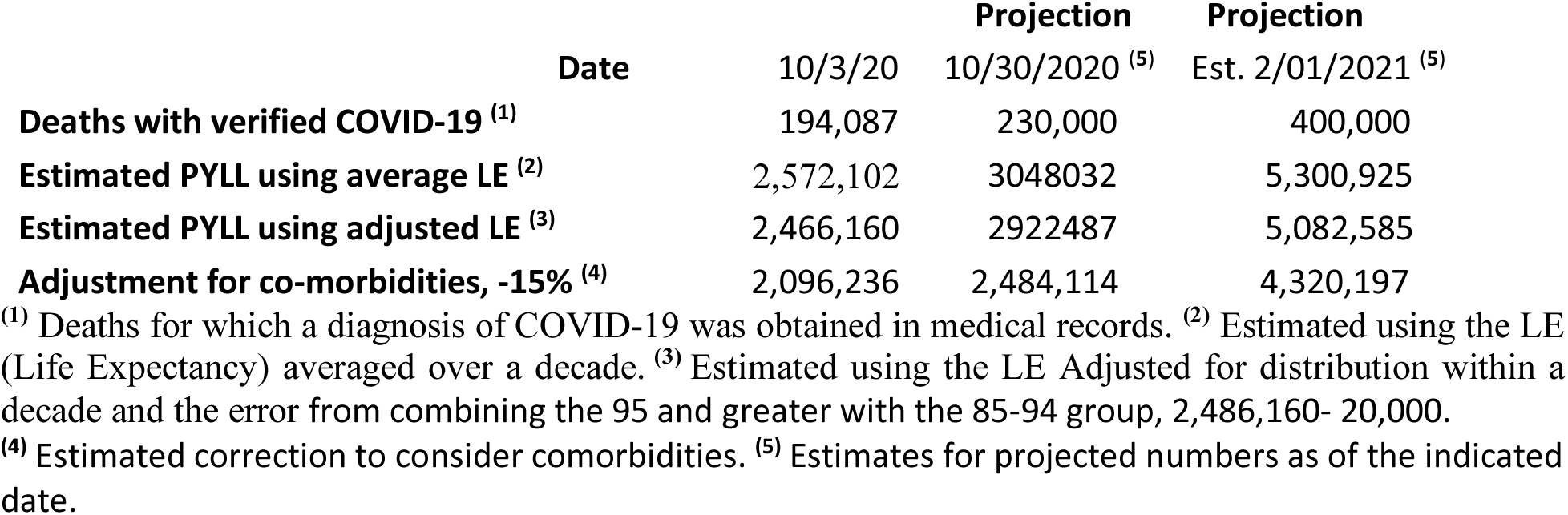
Summary of current and projected person-years of life lost.

One particularly enlightening observation was the relatively uniform rise in the fold increase in COVID-19 deaths per million people from decade to decade of the population. The constant increase may be due to gradual aging leading to a proportional increase in morbidity and mortality. We and others (Montecino-Rodriguez et al., 2013; Shrock et al, 2020) have previously shown that the immune system decreases in efficiency with age. This is also seen in the percent increase in deaths over the normal level after age 45 which is approximately 10% for each decade (Table 2B). This observation together with other aspects of organismal senescence may contribute to this steady increase in susceptibility to COVID-19 lethality with age.

In addition to frank mortality and the person-years of life lost, there exist additional common metrics used for quantifying the overall disease burden and potential advancement of mortality such as disability-adjusted life years (DALYs) (Murray et al., 2012), quality-adjusted life-year (QALY) (Prieto and Sacristán, 2003), and disability-adjusted life expectancy (DALE), among others. We did not undertake those analyses but note that there is a growing awareness of lasting effects on those infected with SARS-CoV-2 that lead to serious medical consequences (Cousin-Frankel, 2020). COVID-19 patients who recover from the infection suffer from a number of potential long-term conditions such as myocarditis, pericarditis, increased propensity for blood clotting and strokes, lung scarring, fatigue, and neurological symptoms like dizziness, loss of taste and smell, impaired consciousness and loss of mental acuity. The long-term ramifications of COVID-19 will become apparent in the fullness of time but it already clear that many survivors suffer from debilitation that may affect the quality of their remaining years of life.

If projections are correct that as many as 400,000 deaths in the US will occur by February 2021 following what is being called the ‘second wave’, then even conservative estimates project that between 4 and 5 million person-years of life will be lost in the United States alone by that time (Table 11). Additional studies have aimed at the possibility that COVID-19 deaths are under-reported. By analyzing what they call “excess deaths” above historical death rates, they estimate that perhaps 50% more deaths can be linked to COVID-19 than directly reported as COVID-19 deaths, many either under-reported COVID-19 deaths or indirect deaths due to stresses on the medical system or reluctance for individuals to seek medical help for underlying conditions (Woolf et al., 2020). Thus, the estimates herein could be expanded to perhaps 600,000 direct and indirect COVID-19 deaths and many additional million person-years of life lost in the next few months in the United States alone. These losses are on the same scale as the 6.8 million person-years lost to cancer in 1984 (Horm and Sondik, 2020). Note that the COVID-19 death toll through April 2021 will represent a single year of mortality.

Converting the COVID-19 death toll from individual deaths to person-years lost distributed across age categories shines a light on the magnitude of the pandemic’s toll across the American population. The loss of 2.5 million person-years might be viewed as the direct cost of the pandemic. COVID-19 has wiped out millions of years of productive, active, and happy existence. One must also consider the indirect costs of these lost person-years in the form of emotional and economic tolls these absences impose on the families, friends and co-workers of those lost. Who among us would not cherish another 5 years together with a father, mother, son, daughter or close friend? The full impact of COVID-19 will emerge over time and it is certain to be enormous.

## Data Availability

All Data used in this study is available in the tables in this paper or the links to the publicly available data.

https://coronavirus.jhu.edu/map.html

https://www.cdc.gov/nchs/nvss/vsrr/covid_weekly/index.htm

https://www.cdc.gov/nchs/nvss/vsrr/COVID19/index.htm

https://www.cdc.gov/coronavirus/2019-ncov/need-extra-precautions/evidence-table.html

https://www.ssa.gov/oact/STATS/table4c6.html#fn1

https://www.census.gov/topics/population/age-and-sex/library/publications.2011.html

https://www.cdc.gov/nchs/nvss/vsrr/covid_weekly/index.htm#Comorbidities

---

**Table 1** Actuarial Life Table for Male and Females in the US in 2017.

**Table 2** COVID-19 Deaths in the US from the CDC as of Oct. 3 2020.

**Table 3** Calculation of the average life expectancy (LE) for indicated age groups.

**Table 4** Calculation of the Potential Years of Life Lost to COVID-19 (person-years) in the US using the life expectancy (LE) averaged over the 10-year periods indicated.

**Table 5** Calculation of base numbers and factors for calculation of deaths monometrically distributed through decades of life categories.

**Table 6** Age distribution adjusted person-years lost calculation.

**Table 7** Population distribution in the US by sex according to the 2000 and 2010 census.

**Table 8** Distribution of COVID-19 associated deaths among different ethnicities.

**Table 9** Actuarial Table for Sex, and Race in the US.

**Table 10** Comorbidity distributions by age groups.

**Table 11** Summary of current and projected person-years of life lost.

## Acknowledgements

We thank D. Wishart, M. Kuroda, M. Murray, C.-C. Huang, H. Zoghbi and J. Glaven for helpful advice, guidance and comments on the manuscript. D. Wishart’s clarifying edits and ability to turn a phrase was tremendously helpful and I am very grateful for his input. S.J.E. thanks D. F. G. for the inspiration that initiated this study when in the course of discussing the then 200,000 deaths due to COVID-19, D.F.G. noted that many say that “they were going to die anyway” so those deaths don’t really matter. This work was supported by grants from the MassCPR and the Value of Vaccine Research Network to S.J.E. S.J.E. is an Investigator with the Howard Hughes Medical Institute.

