## Supplementary material for "2.5 Million Person-Years of Life Have Been Lost Due to COVID-19 in the United States": Table 6 v2

**Period Life Table, 2017**

| Exact age | Male Death Probability (a) | Male Number of Lives (b) | Male Life expectancy | Base number | Factor | Person-Years lost |
|---|---|---|---|---|---|---|
| 0 | 0.006304 | 100,000 | 75.97 | 13 | 1 | 987.61 |
| 1 | 0.000426 | 99,370 | 75.45 | 1.75 | 1 | 132.038 |
| 2 | 0.00029 | 99,327 | 74.48 | 1.75 | 1 | 130.34 |
| 3 | 0.000229 | 99,298 | 73.5 | 1.75 | 1 | 128.625 |
| 4 | 0.000162 | 99,276 | 72.52 | 1.75 | 1 | 126.91 |
| 5 | 0.000146 | 99,260 | 71.53 | 1.438 | 1 | 102.86 |
| 6 | 0.000136 | 99,245 | 70.54 | 1.438 | 1.091 | 110.712 |
| 7 | 0.000127 | 99,232 | 69.55 | 1.438 | 1.191 | 119.14 |
| 8 | 0.000115 | 99,219 | 68.56 | 1.438 | 1.3 | 128.183 |
| 9 | 0.000103 | 99,208 | 67.57 | 1.438 | 1.419 | 137.884 |
| 10 | 0.000097 | 99,197 | 66.57 | 1.438 | 1.549 | 148.265 |
| 11 | 0.000109 | 99,188 | 65.58 | 1.438 | 1.69 | 159.415 |
| 12 | 0.000151 | 99,177 | 64.59 | 1.438 | 1.845 | 171.366 |
| 13 | 0.000232 | 99,162 | 63.6 | 1.438 | 2.014 | 184.169 |
| 14 | 0.000343 | 99,139 | 62.61 | 1.438 | 2.198 | 197.88 |
| 15 | 0.000465 | 99,105 | 61.63 | 14.58 | 1 | 898.424 |
| 16 | 0.000588 | 99,059 | 60.66 | 14.58 | 1.091 | 965.142 |
| 17 | 0.00072 | 99,001 | 59.7 | 14.58 | 1.191 | 1036.72 |
| 18 | 0.000858 | 98,929 | 58.74 | 14.58 | 1.3 | 1113.33 |
| 19 | 0.000999 | 98,845 | 57.79 | 14.58 | 1.419 | 1195.48 |
| 20 | 0.001146 | 98,746 | 56.85 | 14.58 | 1.549 | 1283.57 |
| 21 | 0.001288 | 98,633 | 55.91 | 14.58 | 1.69 | 1377.78 |
| 22 | 0.001407 | 98,506 | 54.98 | 14.58 | 1.845 | 1478.75 |
| 23 | 0.001494 | 98,367 | 54.06 | 14.58 | 2.014 | 1586.96 |
| 24 | 0.001556 | 98,220 | 53.14 | 14.58 | 2.198 | 1702.59 |
| 25 | 0.00161 | 98,067 | 52.22 | 65.57 | 1 | 3423.81 |
| 26 | 0.001665 | 97,910 | 51.31 | 65.57 | 1.091 | 3671.76 |
| 27 | 0.001717 | 97,746 | 50.39 | 65.57 | 1.191 | 3935.66 |
| 28 | 0.001767 | 97,579 | 49.48 | 65.57 | 1.3 | 4217.96 |
| 29 | 0.001817 | 97,406 | 48.56 | 65.57 | 1.419 | 4518.05 |
| 30 | 0.001865 | 97,229 | 47.65 | 65.57 | 1.549 | 4838.78 |
| 31 | 0.001911 | 97,048 | 46.74 | 65.57 | 1.69 | 5180.38 |
| 32 | 0.00196 | 96,862 | 45.83 | 65.57 | 1.845 | 5543.99 |

| | | | | | | |
|---|---|---|---|---|---|---|
| 33 | 0.002014 | 96,672 | 44.92 | 65.57 | 2.014 | 5930.79 |
| 34 | 0.002071 | 96,478 | 44.01 | 65.57 | 2.198 | 6341.97 |
| 35 | 0.002138 | 96,278 | 43.1 | 176.4 | 1 | 7601.36 |
| 36 | 0.002211 | 96,072 | 42.19 | 176.4 | 1.091 | 8121.26 |
| 37 | 0.002279 | 95,860 | 41.28 | 176.4 | 1.191 | 8672.68 |
| 38 | 0.002342 | 95,641 | 40.37 | 176.4 | 1.3 | 9257.05 |
| 39 | 0.002405 | 95,417 | 39.47 | 176.4 | 1.419 | 9878.27 |
| 40 | 0.002482 | 95,188 | 38.56 | 176.4 | 1.549 | 10533 |
| 41 | 0.002583 | 94,951 | 37.65 | 176.4 | 1.69 | 11224.8 |
| 42 | 0.00271 | 94,706 | 36.75 | 176.4 | 1.845 | 11958.3 |
| 43 | 0.00287 | 94,450 | 35.85 | 176.4 | 2.014 | 12732.2 |
| 44 | 0.003064 | 94,178 | 34.95 | 176.4 | 2.198 | 13547.6 |
| 45 | 0.003285 | 93,890 | 34.06 | 461.7 | 1 | 15725.6 |
| 46 | 0.003538 | 93,581 | 33.17 | 461.7 | 1.091 | 16715.1 |
| 47 | 0.003834 | 93,250 | 32.28 | 461.7 | 1.191 | 17754 |
| 48 | 0.004178 | 92,893 | 31.41 | 461.7 | 1.3 | 18855.2 |
| 49 | 0.004569 | 92,505 | 30.54 | 461.7 | 1.419 | 20009.3 |
| 50 | 0.004997 | 92,082 | 29.67 | 461.7 | 1.549 | 21216.8 |
| 51 | 0.005462 | 91,622 | 28.82 | 461.7 | 1.69 | 22493.5 |
| 52 | 0.005971 | 91,122 | 27.98 | 461.7 | 1.845 | 23834.7 |
| 53 | 0.006526 | 90,577 | 27.14 | 461.7 | 2.014 | 25233.2 |
| 54 | 0.007125 | 89,986 | 26.32 | 461.7 | 2.198 | 26708.4 |
| 55 | 0.007766 | 89,345 | 25.5 | 1050 | 1 | 26782.2 |
| 56 | 0.008445 | 88,651 | 24.7 | 1050 | 1.091 | 28314.2 |
| 57 | 0.009156 | 87,903 | 23.9 | 1050 | 1.191 | 29902.3 |
| 58 | 0.009897 | 87,098 | 23.12 | 1050 | 1.3 | 31571.5 |
| 59 | 0.010671 | 86,236 | 22.34 | 1050 | 1.419 | 33295.8 |
| 60 | 0.011519 | 85,316 | 21.58 | 1050 | 1.549 | 35104.1 |
| 61 | 0.012419 | 84,333 | 20.83 | 1050 | 1.69 | 36982.5 |
| 62 | 0.013307 | 83,286 | 20.08 | 1050 | 1.845 | 38910.8 |
| 63 | 0.014164 | 82,177 | 19.35 | 1050 | 2.014 | 40924.9 |
| 64 | 0.015032 | 81,013 | 18.62 | 1050 | 2.198 | 42982 |
| 65 | 0.016013 | 79,795 | 17.89 | 1681 | 1 | 30080.6 |
| 66 | 0.017138 | 78,518 | 17.18 | 1681 | 1.091 | 31528.2 |
| 67 | 0.018362 | 77,172 | 16.47 | 1681 | 1.191 | 32989.1 |
| 68 | 0.019693 | 75,755 | 15.77 | 1681 | 1.3 | 34475.3 |
| 69 | 0.021174 | 74,263 | 15.07 | 1681 | 1.419 | 35957.5 |
| 70 | 0.022889 | 72,691 | 14.39 | 1681 | 1.549 | 37474.6 |
| 71 | 0.024869 | 71,027 | 13.71 | 1681 | 1.69 | 38968.5 |
| 72 | 0.027095 | 69,261 | 13.05 | 1681 | 1.845 | 40484.4 |

| | | | | | | |
|---|---|---|---|---|---|---|
| 73 | 0.029587 | 67,384 | 12.4 | 1681 | 2.014 | 41985.4 |
| 74 | 0.032394 | 65,390 | 11.76 | 1681 | 2.198 | 43459.4 |
| 75 | 0.035668 | 63,272 | 11.14 | 1846 | 1 | 20563.9 |
| 76 | 0.039396 | 61,015 | 10.53 | 1846 | 1.091 | 21215.3 |
| 77 | 0.043453 | 58,611 | 9.94 | 1846 | 1.191 | 21857.9 |
| 78 | 0.047826 | 56,065 | 9.37 | 1846 | 1.3 | 22488.5 |
| 79 | 0.052649 | 53,383 | 8.82 | 1846 | 1.419 | 23104.1 |
| 80 | 0.058206 | 50,573 | 8.28 | 1846 | 1.549 | 23672.9 |
| 81 | 0.064581 | 47,629 | 7.76 | 1846 | 1.69 | 24214.9 |
| 82 | 0.071657 | 44,553 | 7.26 | 1846 | 1.845 | 24726.2 |
| 83 | 0.079465 | 41,361 | 6.79 | 1846 | 2.014 | 25240.1 |
| 84 | 0.088141 | 38,074 | 6.33 | 1846 | 2.198 | 25681.8 |
| 85 | 0.097854 | 34,718 | 5.89 | 1558 | 1 | 9178.58 |
| 86 | 0.108747 | 31,321 | 5.48 | 1558 | 1.091 | 9320.53 |
| 87 | 0.120919 | 27,915 | 5.08 | 1558 | 1.191 | 9430.26 |
| 88 | 0.134425 | 24,539 | 4.71 | 1558 | 1.3 | 9542.91 |
| 89 | 0.149273 | 21,241 | 4.37 | 1558 | 1.419 | 9663.66 |
| 90 | 0.165452 | 18,070 | 4.05 | 1558 | 1.549 | 9774.96 |
| 91 | 0.182935 | 15,080 | 3.75 | 1558 | 1.69 | 9878.51 |
| 92 | 0.201679 | 12,322 | 3.48 | 1558 | 1.845 | 10005.5 |
| 93 | 0.221637 | 9,837 | 3.23 | 1558 | 2.014 | 10135.9 |
| 94 | 0.242747 | 7,656 | 3.01 | 1558 | 2.198 | 10309.2 |
| 95 | 0.263672 | 5,798 | 2.81 | | | **1417459** |
| 96 | 0.284014 | 4,269 | 2.64 | | | |
| 97 | 0.303355 | 3,057 | 2.49 | | | |
| 98 | 0.321268 | 2,129 | 2.36 | | | |
| 99 | 0.337332 | 1,445 | 2.24 | | | |
| 100 | 0.354198 | 958 | 2.12 | | | |
| 101 | 0.371908 | 619 | 2.01 | | | |
| 102 | 0.390503 | 388 | 1.9 | | | |
| 103 | 0.410029 | 237 | 1.8 | | | |
| 104 | 0.43053 | 140 | 1.7 | | | |
| 105 | 0.452057 | 80 | 1.6 | | | |
| 106 | 0.474659 | 44 | 1.51 | | | |
| 107 | 0.498392 | 23 | 1.42 | | | |
| 108 | 0.523312 | 11 | 1.34 | | | |
| 109 | 0.549478 | 5 | 1.26 | | | |
| 110 | 0.576951 | 2 | 1.18 | | | |
| 111 | 0.605799 | 1 | 1.1 | | | |
| 112 | 0.636089 | 0 | 1.03 | | | |

| | | | |
|---|---|---|---|
| 113 | 0.667893 | 0 | 0.96 |
| 114 | 0.701288 | 0 | 0.9 |
| 115 | 0.736353 | 0 | 0.84 |
| 116 | 0.77317 | 0 | 0.78 |
| 117 | 0.811829 | 0 | 0.72 |
| 118 | 0.85242 | 0 | 0.66 |
| 119 | 0.895041 | 0 | 0.61 |

| **Exact age** | **Female Death Probability (a)** | **Female Number of Lives (b)** | **Female Life expectancy** | **Base number** | **Factor** | **Person-Years lost** |
|---|---|---|---|---|---|---|
| 0 | 0.005229 | 100,000 | 80.96 | 7 | 1 | 566.72 |
| 1 | 0.000342 | 99,477 | 80.39 | 2 | 1 | 160.78 |
| 2 | 0.000209 | 99,443 | 79.42 | 2 | 1 | 158.84 |
| 3 | 0.000162 | 99,422 | 78.43 | 2 | 1 | 156.86 |
| 4 | 0.000143 | 99,406 | 77.45 | 2 | 1 | 154.9 |
| 5 | 0.000125 | 99,392 | 76.46 | 0.615 | 1 | 46.9877 |
| 6 | 0.000113 | 99,379 | 75.47 | 0.615 | 1.104 | 51.2182 |
| 7 | 0.000104 | 99,368 | 74.47 | 0.615 | 1.22 | 55.8125 |
| 8 | 0.000097 | 99,358 | 73.48 | 0.615 | 1.347 | 60.8162 |
| 9 | 0.000093 | 99,348 | 72.49 | 0.615 | 1.487 | 66.2565 |
| 10 | 0.000092 | 99,339 | 71.5 | 0.615 | 1.642 | 72.1699 |
| 11 | 0.000098 | 99,330 | 70.5 | 0.615 | 1.814 | 78.5849 |
| 12 | 0.000113 | 99,320 | 69.51 | 0.615 | 2.003 | 85.5652 |
| 13 | 0.000138 | 99,309 | 68.52 | 0.615 | 2.212 | 93.1467 |
| 14 | 0.000172 | 99,295 | 67.53 | 0.615 | 2.443 | 101.379 |
| 15 | 0.000211 | 99,278 | 66.54 | 8.542 | 1 | 568.392 |
| 16 | 0.000251 | 99,257 | 65.55 | 8.542 | 1.104 | 618.355 |
| 17 | 0.000293 | 99,232 | 64.57 | 8.542 | 1.22 | 672.66 |
| 18 | 0.000336 | 99,203 | 63.59 | 8.542 | 1.347 | 731.566 |
| 19 | 0.000379 | 99,170 | 62.61 | 8.542 | 1.487 | 795.442 |
| 20 | 0.000425 | 99,132 | 61.63 | 8.542 | 1.642 | 864.683 |
| 21 | 0.000472 | 99,090 | 60.66 | 8.542 | 1.814 | 939.869 |
| 22 | 0.000515 | 99,044 | 59.69 | 8.542 | 2.003 | 1021.33 |
| 23 | 0.000551 | 98,993 | 58.72 | 8.542 | 2.212 | 1109.56 |
| 24 | 0.000582 | 98,938 | 57.75 | 8.542 | 2.443 | 1205.08 |
| 25 | 0.000612 | 98,880 | 56.78 | 31.34 | 1 | 1779.57 |

| | | | | | | |
|---|---|---|---|---|---|---|
| 26 | 0.000646 | 98,820 | 55.82 | 31.34 | 1.104 | 1932.01 |
| 27 | 0.000684 | 98,756 | 54.85 | 31.34 | 1.22 | 2096.51 |
| 28 | 0.000729 | 98,689 | 53.89 | 31.34 | 1.347 | 2274.72 |
| 29 | 0.000779 | 98,617 | 52.93 | 31.34 | 1.487 | 2467.3 |
| 30 | 0.000833 | 98,540 | 51.97 | 31.34 | 1.642 | 2675.3 |
| 31 | 0.000887 | 98,458 | 51.01 | 31.34 | 1.814 | 2899.85 |
| 32 | 0.000939 | 98,370 | 50.06 | 31.34 | 2.003 | 3142.76 |
| 33 | 0.000988 | 98,278 | 49.1 | 31.34 | 2.212 | 3404.1 |
| 34 | 0.001034 | 98,181 | 48.15 | 31.34 | 2.443 | 3686.52 |
| 35 | 0.001085 | 98,079 | 47.2 | 77.31 | 1 | 3648.99 |
| 36 | 0.001143 | 97,973 | 46.25 | 77.31 | 1.104 | 3948.59 |
| 37 | 0.001205 | 97,861 | 45.3 | 77.31 | 1.22 | 4270.99 |
| 38 | 0.001271 | 97,743 | 44.36 | 77.31 | 1.347 | 4618.73 |
| 39 | 0.001345 | 97,619 | 43.41 | 77.31 | 1.487 | 4991.38 |
| 40 | 0.001429 | 97,488 | 42.47 | 77.31 | 1.642 | 5392.78 |
| 41 | 0.001524 | 97,348 | 41.53 | 77.31 | 1.814 | 5823.62 |
| 42 | 0.00163 | 97,200 | 40.59 | 77.31 | 2.003 | 6285.64 |
| 43 | 0.001748 | 97,042 | 39.66 | 77.31 | 2.212 | 6782.4 |
| 44 | 0.001881 | 96,872 | 38.73 | 77.31 | 2.443 | 7314.39 |
| 45 | 0.002029 | 96,690 | 37.8 | 204.3 | 1 | 7723.85 |
| 46 | 0.002195 | 96,494 | 36.88 | 204.3 | 1.104 | 8322.1 |
| 47 | 0.002386 | 96,282 | 35.96 | 204.3 | 1.22 | 8961.1 |
| 48 | 0.002605 | 96,052 | 35.04 | 204.3 | 1.347 | 9642.86 |
| 49 | 0.002851 | 95,802 | 34.13 | 204.3 | 1.487 | 10372.4 |
| 50 | 0.003118 | 95,529 | 33.23 | 204.3 | 1.642 | 11152.5 |
| 51 | 0.003403 | 95,231 | 32.33 | 204.3 | 1.814 | 11982.5 |
| 52 | 0.003714 | 94,907 | 31.44 | 204.3 | 2.003 | 12868.4 |
| 53 | 0.004052 | 94,554 | 30.55 | 204.3 | 2.212 | 13808.7 |
| 54 | 0.004415 | 94,171 | 29.68 | 204.3 | 2.443 | 14815.1 |
| 55 | 0.004813 | 93,755 | 28.81 | 535 | 1 | 15412.1 |
| 56 | 0.005233 | 93,304 | 27.94 | 535 | 1.104 | 16506.1 |
| 57 | 0.005647 | 92,816 | 27.09 | 535 | 1.22 | 17673.7 |
| 58 | 0.006043 | 92,292 | 26.24 | 535 | 1.347 | 18905.3 |
| 59 | 0.006441 | 91,734 | 25.39 | 535 | 1.487 | 20201.4 |
| 60 | 0.006886 | 91,143 | 24.56 | 535 | 1.642 | 21579.8 |
| 61 | 0.007391 | 90,515 | 23.72 | 535 | 1.814 | 23016.2 |
| 62 | 0.007931 | 89,846 | 22.9 | 535 | 2.003 | 24538.9 |
| 63 | 0.008508 | 89,134 | 22.07 | 535 | 2.212 | 26116.9 |
| 64 | 0.009142 | 88,375 | 21.26 | 535 | 2.443 | 27783.2 |
| 65 | 0.009874 | 87,568 | 20.45 | 992.1 | 1 | 20287.5 |

| | | | | | | |
|---|---|---|---|---|---|---|
| 66 | 0.010717 | 86,703 | 19.65 | 992.1 | 1.104 | 21527.7 |
| 67 | 0.01166 | 85,774 | 18.86 | 992.1 | 1.22 | 22817.9 |
| 68 | 0.012711 | 84,774 | 18.07 | 992.1 | 1.347 | 24143.1 |
| 69 | 0.013894 | 83,696 | 17.3 | 992.1 | 1.487 | 25525.9 |
| 70 | 0.015285 | 82,533 | 16.54 | 992.1 | 1.642 | 26950.7 |
| 71 | 0.016878 | 81,272 | 15.79 | 992.1 | 1.814 | 28413 |
| 72 | 0.018607 | 79,900 | 15.05 | 992.1 | 2.003 | 29906.9 |
| 73 | 0.020466 | 78,413 | 14.32 | 992.1 | 2.212 | 31425.2 |
| 74 | 0.022522 | 76,809 | 13.61 | 992.1 | 2.443 | 32983.2 |
| 75 | 0.024929 | 75,079 | 12.92 | 1421 | 1 | 18360.9 |
| 76 | 0.027729 | 73,207 | 12.23 | 1421 | 1.104 | 19193.7 |
| 77 | 0.030855 | 71,177 | 11.57 | 1421 | 1.22 | 20052.3 |
| 78 | 0.034321 | 68,981 | 10.92 | 1421 | 1.347 | 20900.4 |
| 79 | 0.038211 | 66,613 | 10.29 | 1421 | 1.487 | 21749.4 |
| 80 | 0.042771 | 64,068 | 9.68 | 1421 | 1.642 | 22594.7 |
| 81 | 0.047992 | 61,328 | 9.09 | 1421 | 1.814 | 23431.3 |
| 82 | 0.053678 | 58,385 | 8.52 | 1421 | 2.003 | 24253.3 |
| 83 | 0.05981 | 55,251 | 7.98 | 1421 | 2.212 | 25086.2 |
| 84 | 0.066584 | 51,946 | 7.45 | 1421 | 2.443 | 25863.6 |
| 85 | 0.074258 | 48,487 | 6.95 | 2210 | 1 | 15359.1 |
| 86 | 0.083053 | 44,887 | 6.47 | 2210 | 1.104 | 15790.1 |
| 87 | 0.093123 | 41,159 | 6.01 | 2210 | 1.22 | 16197.8 |
| 88 | 0.10454 | 37,326 | 5.57 | 2210 | 1.347 | 16578.2 |
| 89 | 0.117305 | 33,424 | 5.16 | 2210 | 1.487 | 16960.2 |
| 90 | 0.131392 | 29,503 | 4.78 | 2210 | 1.642 | 17350.4 |
| 91 | 0.146753 | 25,627 | 4.43 | 2210 | 1.814 | 17757.7 |
| 92 | 0.163331 | 21,866 | 4.11 | 2210 | 2.003 | 18193.8 |
| 93 | 0.181064 | 18,294 | 3.81 | 2210 | 2.212 | 18625.5 |
| 94 | 0.199886 | 14,982 | 3.55 | 2210 | 2.443 | 19165.1 |
| 95 | 0.218908 | 11,987 | 3.31 | | | **1068701** |
| 96 | 0.237815 | 9,363 | 3.09 | | | |
| 97 | 0.256265 | 7,136 | 2.9 | | | |
| 98 | 0.273894 | 5,308 | 2.73 | | | |
| 99 | 0.290328 | 3,854 | 2.58 | | | |
| 100 | 0.307747 | 2,735 | 2.42 | | | |
| 101 | 0.326212 | 1,893 | 2.28 | | | |
| 102 | 0.345785 | 1,276 | 2.14 | | | |
| 103 | 0.366532 | 835 | 2.01 | | | |
| 104 | 0.388524 | 529 | 1.88 | | | |
| 105 | 0.411835 | 323 | 1.76 | | | |

| | | | |
|---|---|---|---|
| 106 | 0.436546 | 190 | 1.65 |
| 107 | 0.462738 | 107 | 1.54 |
| 108 | 0.490503 | 58 | 1.44 |
| 109 | 0.519933 | 29 | 1.34 |
| 110 | 0.551129 | 14 | 1.24 |
| 111 | 0.584196 | 6 | 1.15 |
| 112 | 0.619248 | 3 | 1.06 |
| 113 | 0.656403 | 1 | 0.98 |
| 114 | 0.695787 | 0 | 0.91 |
| 115 | 0.736353 | 0 | 0.84 |
| 116 | 0.77317 | 0 | 0.78 |
| 117 | 0.811829 | 0 | 0.72 |
| 118 | 0.85242 | 0 | 0.66 |
| 119 | 0.895041 | 0 | 0.61 |

Note: The period life expectancy at a given age for 2017 represents the average number of years of life remaining if a group of persons at that age were to experience the mortality rates for 2017 over the course of their remaining life.

[a] Probability of dying within one year.

[b] Number of survivors out of 100,000 born alive.

[c] Base numbers represent are uniform throughout the period of years in each category and are derived such as the sum of the produce of the base number and the factor are equal to the total number of deaths in that span of years in total. For time spans of less that 10 years, such as 1-4 years, the number of deaths is evenly divided among the 4 years and the base number is 1. For the category of greater than 85 years all remaining in deaths are assumed to be n the 85-94 years span and all deaths and the base factors are adjusted to that. This slightly over estimate the person years became some of deaths occur at age 95 and older but this is a small number and estimated an extra 1-2 years of life expectancy and therefor adds a relatively small number of years. We estimate only a total of 15,000 COVID deaths are after the age of 95 and therefore approximately 10-20,000 extra person years may be artificially added to the final total. See discussion in Table 4.
