## Supplementary material for "2.5 Million Person-Years of Life Have Been Lost Due to COVID-19 in the United States": Table 5

**Table 5. Calculation of base numbers and factors for calculation of deaths monometrically distributed through decades of life categories**

**Table 5A**

**Females**

| All ages | Total Deaths in age group | Total Population | Deaths per million | Fold more likely to die in the next decade from COVID. | Base number for distribution of deaths per year |
|---|---|---|---|---|---|
| Under 1 year | 7 | 1,879,703 | 3.7 | | |
| 1–4 years | 8 | 7,798,370 | 1 | | |
| 5–14 years | 10 | 20,100,339 | 0.5 | | 0.614539962 |
| 15–24 years | 139 | 20,994,345 | 6.6 | **3.44** | 8.542105474 |
| 25–34 years | 510 | 22,487,065 | 22.7 | **2.68** | 31.34153807 |
| 35–44 years | 1,258 | 20,690,288 | 60.8 | **2.59** | 77.30912724 |
| 45–54 years | 3,325 | 21,090,497 | 157.7 | **2.52** | 204.3345374 |
| 55–64 years | 8,705 | 21,873,773 | 398 | **2.5** | 534.9570371 |
| 65–74 years | 16,143 | 16,246,231 | 994 | **2.69** | 992.0518609 |
| 75–84 years | 23,125 | 8,659,334 | 2671 | **3.19** | 1421.123663 |
| 85 years and | 35,961 | 4,218,810 | 8524 | | 2209.947158 |
| | 89191 | | | **AVE = 2.698 from 35-84** | |
| | | | | The log10 of 2.698 = 0.431 | |
| | | | | The log10 of 1.10433 = 0.0431 | |

**Males**

| All ages | Total Deaths in age group | Total Population | Deaths per million | likely to die in the next decade from COVID. | Base number for distributio n of deaths per year |
|---|---|---|---|---|---|
| Under 1 year | 13 | 1,968,505 | | | |
| 1–4 years | 7 | 8,163,697 | 0.35 | | |
| 5–14 years | 22 | 20,974,830 | 1 | | 1.4381184 |
| 15–24 years | 223 | 21,976,455 | 10 | **4.4** | 14.577291 |
| 25–34 years | 1,003 | 23,210,709 | 43 | **3** | 65.565127 |
| 35–44 years | 2,698 | 20,587,600 | 131 | **2.63** | 176.36561 |
| 45–54 years | 7,063 | 20,541,202 | 344 | **2.3** | 461.70139 |
| 55–64 years | 16,067 | 20,398,863 | 788 | **2.3** | 1050.284 |
| 65–74 years | 25,722 | 14,246,085 | 1806 | **2.3** | 1681.4219 |
| 75–84 years | 28,239 | 6,735,040 | 4193 | **2.45** | 1845.9557 |
| 85 years and ove | 23,839 | 2,325,693 | 10253 | | 1558.3321 |
| | 104896 | | | **AVE = 2.396 from 35-84** | |
| | | | | The log10 of 2.396 = 0.380 | |
| | | | | The log10 of 1.09144 = 0.038 | |

**Deaths per decade formula** $\sum_{i=0}^{9} A(1+r)^i$

**Table 5B Calculation of the (1 +r) variable**

| Year in serie | Female factors (1 +r) | exponent | Male factors (1 | exponent | |
|---|---|---|---|---|---|
| 1 | 1 | 0 | 1 | 0 | 1.00E+00 |
| 2 | 1.104332873 | 1 | 1.091440336 | 1 | 1e0.038 |
| 3 | 1.219551094 | 2 | 1.191242008 | 2 | 1e0.076 |
| 4 | 1.346790363 | 3 | 1.300169578 | 3 | 1e0.114 |
| 5 | 1.48730487 | 4 | 1.419057522 | 4 | 1e0.152 |
| 6 | 1.64247966 | 5 | 1.548816619 | 5 | 1e0.190 |
| 7 | 1.813844281 | 6 | 1.690440932 | 6 | 1e0.228 |
| 8 | 2.003087865 | 7 | 1.845015419 | 7 | 1e0.266 |
| 9 | 2.212075776 | 8 | 2.01372425 | 8 | 1e0.304 |
| 10 | 2.442867997 | 9 | 2.197859873 | 9 | 1e0.342 |
| SUM | 16.27233478 | | 15.29776654 | | |

### Descriptions of the calculations

(a) The male and female factors were calculated by measuring fold increase in average COVID-19 deaths per million from one decade to the next and taking the average fold increase for the decade spanning 35-44 to the decade spanning 75-84. The decades 15-24 and 25-34 were left out of the calculation as the total deaths were low and the fold increase became relatively stable after those years. A second source of error derives from the assumption that the individuals in the 10-year groups are evenly distributed for deaths. In the absence of smaller grouping, it is not possible to definitively overcome this at this time.

(b) For age groupings other than 10 years the base number was calculated by evenly dividing the total deaths by the number of years the group covered. For the 1-4 group the total deaths were divided by 4 and the factor used was 1.

(c) For 85 and greater, the total deaths were summed and treated as if they were all in the 85-94 range and is further discussed in the text and in Table 4.

| | | | | | | | | |
|---|---|---|---|---|---|---|---|---|
| 43 | **Table 5C Base number (A) calculation** | | | | | | | |
| 44 | | **Female** | | | | **Male** | | |
| 45 | | **All ages** | **Total Deaths in age group** | **Base Number (A)** | | **All ages** | **Total Deaths in age group** | **Base Number (A)** |
| 46 | | Under 1 year | 7 | 7 | | Under 1 year | 13 | 13 |
| 47 | | 1–4 years | 8 | 2 | | 1–4 years | 7 | 1.75 |
| 48 | | 5–14 years | 10 | 0.61454 | | 5–14 years | 22 | 1.4381184 |
| 49 | | 15–24 years | 139 | 8.5421055 | | 15–24 years | 223 | 14.577291 |
| 50 | | 25–34 years | 510 | 31.341538 | | 25–34 years | 1,003 | 65.565127 |
| 51 | | 35–44 years | 1,258 | 77.309127 | | 35–44 years | 2,698 | 176.36561 |
| 52 | | 45–54 years | 3,325 | 204.33454 | | 45–54 years | 7,063 | 461.70139 |
| 53 | | 55–64 years | 8,705 | 534.95704 | | 55–64 years | 16,067 | 1050.284 |
| 54 | | 65–74 years | 16,143 | 992.05186 | | 65–74 years | 25,722 | 1681.4219 |
| 55 | | 75–84 years | 23,125 | 1421.1237 | | 75–84 years | 28,239 | 1845.9557 |
| 56 | | 85 years and | 35,961 | 2209.9472 | | 85 years and over | 23,839 | 1558.3321 |
| 57 | | | | | | | 104896 | |
| 58 | | | | | | | | |
| 59 | | | | | | | | |
