## Supplementary material for "2.5 Million Person-Years of Life Have Been Lost Due to COVID-19 in the United States": Table 4

**Table 4. Calculation of the Potential Years of Life Lost to COVID-19 (person-years)in the US using the  life expectancy (LE) averaged over the 10 year periods indicated**

| | | | |
|---|---|---|---|
| **Males** | | | |
| All ages | 104,896 | Average LE | Person-Years Lost |
| Under 1 year | 13 | 75.97 | 987.61 |
| 1–4 years | 7 | 73.9875 | 517.9125 |
| 5–14 years | 22 | 67.07 | 1475.54 |
| 15–24 years | 223 | 57.346 | 12788.158 |
| 25–34 years | 1,003 | 48.111 | 48255.333 |
| 35–44 years | 2,698 | 39.017 | 105267.87 |
| 45–54 years | 7,063 | 30.139 | 212871.76 |
| 55–64 years | 16,067 | 22.002 | 353506.13 |
| 65–74 years | 25,722 | 14.769 | 379888.22 |
| 75–84 years | 28,239 | 8.622 | 243476.66 |
| 85 years and over (a) | 23,839 | 4.305 | 102626.9 |
| Total | | | 1461662.1 |
| **Females** | | | |
| All ages | 89,191 | Average LE | Person-Years Lost |
| Under 1 year | 7 | 80.96 | 566.72 |
| 1–4 years | 8 | 78.9225 | 631.38 |
| 5–14 years | 10 | 71.993 | 719.93 |
| 15–24 years | 139 | 62.131 | 8636.21 |
| 25–34 years | 510 | 52.456 | 26752.5 |
| 35–44 years | 1,258 | 42.95 | 54031.1 |
| 45–54 years | 3,325 | 33.704 | 112065.8 |
| 55–64 years | 8,705 | 24.998 | 217607.59 |
| 65–74 years | 16,143 | 16.964 | 273849.85 |
| 75–84 years | 23,125 | 10.065 | 232753.13 |
| 85 years and over (a) | 35,961 | 5.084 | 182825.72 |
| Total | | | 1110439.9 |
| Toal person-years Lost | | 2,572,102 ( 1,461662 Males and 1,110,440 Females) | |
| Average person years lost per individual | | | 13.25 years |
| | | | 13.93 years per male |
| | | | 12.45 years per female |

### Potential sources of error in these calculations

(a) A possible source of error results from the fact that the >85 category presented by the CDC did not break down the number of different years. We took the average life expectancy for the 10-year period 85-94 for the entire category. However, of the population of all individuals 85 and older, only 5.1% on average of men are 95 or older and 7.2% of women with average life expectancies of 2.2 and 2.5 years, respectively (See Table 7). There are 1200 men and 2600 women in the 95 and greater category who were originally evaluated at 4 and 5 years of life respectively when the real numbers are closer to 2.2 and 2.5 years, respectively, meaning we overestimated men by 1.8 years and women by 2.5 years of life expectancy. Altogether this is an error of less than 10,000 person years assuming the two groups die at an equal rate. If the 95 and greater dies at a 2-fold rate relative to the 85-94 group, that would be adjusted to 20,000 person-years.

(b) A second source of error derives from the assumption that the individuals in the 10 year groups are evenly distributed for deaths. In the absence of smaller grouping, it is not possible to definitively overcome this at this time.

(c) A third source of error is the inability to account for comorbidities for which we would need the breakdown of deaths per age group for the subgroups for comorbidities and the average life expectancy tables for each comorbidity without COVID-19.
