## Supplementary material for "2.5 Million Person-Years of Life Have Been Lost Due to COVID-19 in the United States": Table 3

| | A | B | C | D | E | F | G | H | I | J | K | L |
|---|---|---|---|---|---|---|---|---|---|---|---|---|
| 1 | **Table 3. Calculation of the average life expectancy (LE) for the indicated age groups** | | | | | | | | | | | |
| 2 | | | | | | | | | | | | |
| 3 | **Life expectancy of Males over the indicated span of age** | | | | | | | | | | | |
| 4 | **Age Span** | 0-1 | 1 to 4 | 5 to 14 | 15-24 | 25-34 | 35-44 | 45-54 | 55-64 | 65-74 | 75-84 | 85-94 |
| 5 | | | 75.45 | 71.53 | 61.63 | 52.22 | 43.1 | 34.06 | 25.5 | 17.89 | 11.14 | 5.89 |
| 6 | | | 74.48 | 70.54 | 60.66 | 51.31 | 42.19 | 33.17 | 24.7 | 17.18 | 10.53 | 5.48 |
| 7 | | | 73.5 | 69.55 | 59.7 | 50.39 | 41.28 | 32.28 | 23.9 | 16.47 | 9.94 | 5.08 |
| 8 | | | 72.52 | 68.56 | 58.74 | 49.48 | 40.37 | 31.41 | 23.12 | 15.77 | 9.37 | 4.71 |
| 9 | | | | 67.57 | 57.79 | 48.56 | 39.47 | 30.54 | 22.34 | 15.07 | 8.82 | 4.37 |
| 10 | | | | 66.57 | 56.85 | 47.65 | 38.56 | 29.67 | 21.58 | 14.39 | 8.28 | 4.05 |
| 11 | | | | 65.58 | 55.91 | 46.74 | 37.65 | 28.82 | 20.83 | 13.71 | 7.76 | 3.75 |
| 12 | | | | 64.59 | 54.98 | 45.83 | 36.75 | 27.98 | 20.08 | 13.05 | 7.26 | 3.48 |
| 13 | | | | 63.6 | 54.06 | 44.92 | 35.85 | 27.14 | 19.35 | 12.4 | 6.79 | 3.23 |
| 14 | | | | 62.61 | 53.14 | 44.01 | 34.95 | 26.32 | 18.62 | 11.76 | 6.33 | 3.01 |
| 15 | Average LE in years | 75.97 | 73.9875 | 67.07 | 57.346 | 48.111 | 39.017 | 30.139 | 22.002 | 14.769 | 8.622 | 4.305 |
| 16 | | | | | | | | | | | | |
| 17 | | | | | | | | | | | | |
| 18 | | | | | | | | | | | | |
| 19 | **Life expectancy of Females over the indicated span of age** | | | | | | | | | | | |
| 20 | **Age Span** | 0-1 | 1 to 4 | 5 to 14 | 15-24 | 25-34 | 35-44 | 45-54 | 55-64 | 65-74 | 75-84 | 85-94 |
| 21 | | | 80.39 | 76.46 | 66.54 | 56.78 | 47.2 | 37.8 | 28.81 | 20.45 | 12.92 | 6.95 |
| 22 | | | 79.42 | 75.47 | 65.55 | 55.82 | 46.25 | 36.88 | 27.94 | 19.65 | 12.23 | 6.47 |
| 23 | | | 78.43 | 74.47 | 64.57 | 54.85 | 45.3 | 35.96 | 27.09 | 18.86 | 11.57 | 6.01 |
| 24 | | | 77.45 | 73.48 | 63.59 | 53.89 | 44.36 | 35.04 | 26.24 | 18.07 | 10.92 | 5.57 |
| 25 | | | 78.9225 | 72.49 | 62.61 | 52.93 | 43.41 | 34.13 | 25.39 | 17.3 | 10.29 | 5.16 |
| 26 | | | | 71.5 | 61.63 | 51.97 | 42.47 | 33.23 | 24.56 | 16.54 | 9.68 | 4.78 |
| 27 | | | | 70.5 | 60.66 | 51.01 | 41.53 | 32.33 | 23.72 | 15.79 | 9.09 | 4.43 |
| 28 | | | | 69.51 | 59.69 | 50.06 | 40.59 | 31.44 | 22.9 | 15.05 | 8.52 | 4.11 |
| 29 | | | | 68.52 | 58.72 | 49.1 | 39.66 | 30.55 | 22.07 | 14.32 | 7.98 | 3.81 |
| 30 | | | | 67.53 | 57.75 | 48.15 | 38.73 | 29.68 | 21.26 | 13.61 | 7.45 | 3.55 |
| 31 | Average LE in years | 80.96 | 78.9225 | 71.993 | 62.131 | 52.456 | 42.95 | 33.704 | 24.998 | 16.964 | 10.065 | 5.084 |
