## Supplementary material for "2.5 Million Person-Years of Life Have Been Lost Due to COVID-19 in the United States": Table 2

| | A | B | C | D | E | F | G | H | I | J | K |
|---|---|---|---|---|---|---|---|---|---|---|---|
| 1 | Table 2. COVID-19 Deaths in the US from the CDC as of OCT. 3 2020 https://www.cdc.gov/nchs/nvss/vsrr/covid_weekly/index.htm |  |  |  |  |  |  |  |  |  |  |
| 2 | **2A Deaths by Gender** | | | | | | | | | | |
| 3 | **Males** | | **COVID Deaths** | **All deaths** | **Population** | | **Females** | | **COVID Deaths** | **All deaths** | **Population** |
| 4 | | All ages | 104,896 | 1,089,340 | 161,128,679 | | | All ages | 89,191 | 998,082 | 166,038,755 |
| 5 | | Under 1 year | 13 | 6,487 | 1,968,505 | | | Under 1 year | 7 | 5,212 | 1,847,935 |
| 6 | | 1–4 years | 7 | 1,305 | 8,163,697 | | | 1–4 years | 8 | 928 | 7,798,370 |
| 7 | | 5–14 years | 22 | 2,062 | 20,974,830 | | | 5–14 years | 10 | 1,414 | 20,100,339 |
| 8 | | 15–24 years | 223 | 16,595 | 21,976,455 | | | 15–24 years | 139 | 5,789 | 20,994,345 |
| 9 | | 25–34 years | 1,003 | 32,519 | 23,210,709 | | | 25–34 years | 510 | 13,512 | 22,487,065 |
| 10 | | 35–44 years | 2,698 | 42,918 | 20,587,600 | | | 35–44 years | 1,258 | 22,136 | 20,690,288 |
| 11 | | 45–54 years | 7,063 | 75,065 | 20,541,202 | | | 45–54 years | 3,325 | 44,368 | 21,090,497 |
| 12 | | 55–64 years | 16,067 | 168,282 | 20,398,863 | | | 55–64 years | 8,705 | 104,901 | 21,873,773 |
| 13 | | 65–74 years | 25,722 | 240,467 | 14,246,085 | | | 65–74 years | 16,143 | 173,389 | 16,246,231 |
| 14 | | 75–84 years | 28,239 | 262,137 | 6,735,040 | | | 75–84 years | 23,125 | 242,282 | 8,659,334 |
| 15 | | 85 years and | 23,839 | 241,503 | 2,325,693 | | | 85 years and | 35,961 | 384,151 | 4,218,810 |
| 16 | | | | | | | | | | | |
| 17 | **2B Percent increased probability of dying relative to non-COVID-19 deaths** | | | | | | | | | | |
| 18 | | | | | | | | | | | |
| 19 | **Sex** | **Age group** | | **194,087** | **Deaths from** | **Population** | | | | | |
| 20 | Total | | | | | Deaths not involving COVID-19 | Increased Pr | Percent Increased probability | | | |
| 21 | | All ages | 203,043 | 2,203,637 | 328,239,523 | 2,000,594 | 0.10149136 | 10.15% | | | |
| 22 | | Under 1 year | 22 | 12,405 | 3,783,052 | 12,383 | 0.00177663 | 0.18% | | | |
| 23 | | 1–4 years | 15 | 2,351 | 15,793,631 | 2,336 | 0.00642123 | 0.64% | | | |
| 24 | | 5–14 years | 37 | 3,690 | 40,994,163 | 3,653 | 0.01012866 | 1.01% | | | |
| 25 | | 15–24 years | 374 | 23,838 | 42,687,510 | 23,464 | 0.01593931 | 1.59% | | | |
| 26 | | 25–34 years | 1,588 | 48,782 | 45,940,321 | 47,194 | 0.03364835 | 3.36% | | | |
| 27 | | 35–44 years | 4,119 | 68,966 | 41,659,144 | 64,847 | 0.06351874 | 6.35% | | | |
| 28 | | 45–54 years | 10,837 | 126,245 | 40,874,902 | 115,408 | 0.09390164 | 9.39% | | | |
| 29 | | 55–64 years | 25,971 | 288,746 | 42,448,537 | 262,775 | 0.0988336 | 9.88% | | | |
| 30 | | 65–74 years | 43,927 | 437,155 | 31,483,433 | 393,228 | 0.11170873 | 11.17% | | | |
| 31 | | 75–84 years | 53,796 | 532,277 | 15,969,872 | 478,481 | 0.1124308 | 11.24% | | | |
| 32 | | 85 years and | 62,357 | 659,182 | 6,604,958 | 596,825 | 0.10448121 | 10.45% | | | |
| 33 | | | | | | | | | | | |
| 34 | | 1 Data from Oct. 7, 2020 | | | | | | | | | |
