## Supplementary material for "2.5 Million Person-Years of Life Have Been Lost Due to COVID-19 in the United States": Table 1

**Table 1. Actuarial Life Table for Male and Females in the US in 2017**
**https://www.ssa.gov/oact/STATS/table4c6.html#fn1**
**Period Life Table, 2017**

| Exact age | Male Death Probability (a) | Male Number of Lives (b) | Male Life expectancy | Female Death Probability (a) | Female Number of Lives (b) | Female Life expectancy |
|---|---|---|---|---|---|---|
| 0 | 0.006304 | 100,000 | 75.97 | 0.005229 | 100,000 | 80.96 |
| 1 | 0.000426 | 99,370 | 75.45 | 0.000342 | 99,477 | 80.39 |
| 2 | 0.00029 | 99,327 | 74.48 | 0.000209 | 99,443 | 79.42 |
| 3 | 0.000229 | 99,298 | 73.5 | 0.000162 | 99,422 | 78.43 |
| 4 | 0.000162 | 99,276 | 72.52 | 0.000143 | 99,406 | 77.45 |
| 5 | 0.000146 | 99,260 | 71.53 | 0.000125 | 99,392 | 76.46 |
| 6 | 0.000136 | 99,245 | 70.54 | 0.000113 | 99,379 | 75.47 |
| 7 | 0.000127 | 99,232 | 69.55 | 0.000104 | 99,368 | 74.47 |
| 8 | 0.000115 | 99,219 | 68.56 | 0.000097 | 99,358 | 73.48 |
| 9 | 0.000103 | 99,208 | 67.57 | 0.000093 | 99,348 | 72.49 |
| 10 | 0.000097 | 99,197 | 66.57 | 0.000092 | 99,339 | 71.5 |
| 11 | 0.000109 | 99,188 | 65.58 | 0.000098 | 99,330 | 70.5 |
| 12 | 0.000151 | 99,177 | 64.59 | 0.000113 | 99,320 | 69.51 |
| 13 | 0.000232 | 99,162 | 63.6 | 0.000138 | 99,309 | 68.52 |
| 14 | 0.000343 | 99,139 | 62.61 | 0.000172 | 99,295 | 67.53 |
| 15 | 0.000465 | 99,105 | 61.63 | 0.000211 | 99,278 | 66.54 |
| 16 | 0.000588 | 99,059 | 60.66 | 0.000251 | 99,257 | 65.55 |
| 17 | 0.00072 | 99,001 | 59.7 | 0.000293 | 99,232 | 64.57 |
| 18 | 0.000858 | 98,929 | 58.74 | 0.000336 | 99,203 | 63.59 |
| 19 | 0.000999 | 98,845 | 57.79 | 0.000379 | 99,170 | 62.61 |
| 20 | 0.001146 | 98,746 | 56.85 | 0.000425 | 99,132 | 61.63 |
| 21 | 0.001288 | 98,633 | 55.91 | 0.000472 | 99,090 | 60.66 |
| 22 | 0.001407 | 98,506 | 54.98 | 0.000515 | 99,044 | 59.69 |
| 23 | 0.001494 | 98,367 | 54.06 | 0.000551 | 98,993 | 58.72 |
| 24 | 0.001556 | 98,220 | 53.14 | 0.000582 | 98,938 | 57.75 |
| 25 | 0.00161 | 98,067 | 52.22 | 0.000612 | 98,880 | 56.78 |
| 26 | 0.001665 | 97,910 | 51.31 | 0.000646 | 98,820 | 55.82 |
| 27 | 0.001717 | 97,746 | 50.39 | 0.000684 | 98,756 | 54.85 |
| 28 | 0.001767 | 97,579 | 49.48 | 0.000729 | 98,689 | 53.89 |
| 29 | 0.001817 | 97,406 | 48.56 | 0.000779 | 98,617 | 52.93 |
| 30 | 0.001865 | 97,229 | 47.65 | 0.000833 | 98,540 | 51.97 |
| 31 | 0.001911 | 97,048 | 46.74 | 0.000887 | 98,458 | 51.01 |
| 32 | 0.00196 | 96,862 | 45.83 | 0.000939 | 98,370 | 50.06 |

| | | | | | | |
|---|---|---|---|---|---|---|
| 33 | 0.002014 | 96,672 | 44.92 | 0.000988 | 98,278 | 49.1 |
| 34 | 0.002071 | 96,478 | 44.01 | 0.001034 | 98,181 | 48.15 |
| 35 | 0.002138 | 96,278 | 43.1 | 0.001085 | 98,079 | 47.2 |
| 36 | 0.002211 | 96,072 | 42.19 | 0.001143 | 97,973 | 46.25 |
| 37 | 0.002279 | 95,860 | 41.28 | 0.001205 | 97,861 | 45.3 |
| 38 | 0.002342 | 95,641 | 40.37 | 0.001271 | 97,743 | 44.36 |
| 39 | 0.002405 | 95,417 | 39.47 | 0.001345 | 97,619 | 43.41 |
| 40 | 0.002482 | 95,188 | 38.56 | 0.001429 | 97,488 | 42.47 |
| 41 | 0.002583 | 94,951 | 37.65 | 0.001524 | 97,348 | 41.53 |
| 42 | 0.00271 | 94,706 | 36.75 | 0.00163 | 97,200 | 40.59 |
| 43 | 0.00287 | 94,450 | 35.85 | 0.001748 | 97,042 | 39.66 |
| 44 | 0.003064 | 94,178 | 34.95 | 0.001881 | 96,872 | 38.73 |
| 45 | 0.003285 | 93,890 | 34.06 | 0.002029 | 96,690 | 37.8 |
| 46 | 0.003538 | 93,581 | 33.17 | 0.002195 | 96,494 | 36.88 |
| 47 | 0.003834 | 93,250 | 32.28 | 0.002386 | 96,282 | 35.96 |
| 48 | 0.004178 | 92,893 | 31.41 | 0.002605 | 96,052 | 35.04 |
| 49 | 0.004569 | 92,505 | 30.54 | 0.002851 | 95,802 | 34.13 |
| 50 | 0.004997 | 92,082 | 29.67 | 0.003118 | 95,529 | 33.23 |
| 51 | 0.005462 | 91,622 | 28.82 | 0.003403 | 95,231 | 32.33 |
| 52 | 0.005971 | 91,122 | 27.98 | 0.003714 | 94,907 | 31.44 |
| 53 | 0.006526 | 90,577 | 27.14 | 0.004052 | 94,554 | 30.55 |
| 54 | 0.007125 | 89,986 | 26.32 | 0.004415 | 94,171 | 29.68 |
| 55 | 0.007766 | 89,345 | 25.5 | 0.004813 | 93,755 | 28.81 |
| 56 | 0.008445 | 88,651 | 24.7 | 0.005233 | 93,304 | 27.94 |
| 57 | 0.009156 | 87,903 | 23.9 | 0.005647 | 92,816 | 27.09 |
| 58 | 0.009897 | 87,098 | 23.12 | 0.006043 | 92,292 | 26.24 |
| 59 | 0.010671 | 86,236 | 22.34 | 0.006441 | 91,734 | 25.39 |
| 60 | 0.011519 | 85,316 | 21.58 | 0.006886 | 91,143 | 24.56 |
| 61 | 0.012419 | 84,333 | 20.83 | 0.007391 | 90,515 | 23.72 |
| 62 | 0.013307 | 83,286 | 20.08 | 0.007931 | 89,846 | 22.9 |
| 63 | 0.014164 | 82,177 | 19.35 | 0.008508 | 89,134 | 22.07 |
| 64 | 0.015032 | 81,013 | 18.62 | 0.009142 | 88,375 | 21.26 |
| 65 | 0.016013 | 79,795 | 17.89 | 0.009874 | 87,568 | 20.45 |
| 66 | 0.017138 | 78,518 | 17.18 | 0.010717 | 86,703 | 19.65 |
| 67 | 0.018362 | 77,172 | 16.47 | 0.01166 | 85,774 | 18.86 |
| 68 | 0.019693 | 75,755 | 15.77 | 0.012711 | 84,774 | 18.07 |
| 69 | 0.021174 | 74,263 | 15.07 | 0.013894 | 83,696 | 17.3 |
| 70 | 0.022889 | 72,691 | 14.39 | 0.015285 | 82,533 | 16.54 |
| 71 | 0.024869 | 71,027 | 13.71 | 0.016878 | 81,272 | 15.79 |
| 72 | 0.027095 | 69,261 | 13.05 | 0.018607 | 79,900 | 15.05 |

| | | | | | | |
|---|---|---|---|---|---|---|
| 73 | 0.029587 | 67,384 | 12.4 | 0.020466 | 78,413 | 14.32 |
| 74 | 0.032394 | 65,390 | 11.76 | 0.022522 | 76,809 | 13.61 |
| 75 | 0.035668 | 63,272 | 11.14 | 0.024929 | 75,079 | 12.92 |
| 76 | 0.039396 | 61,015 | 10.53 | 0.027729 | 73,207 | 12.23 |
| 77 | 0.043453 | 58,611 | 9.94 | 0.030855 | 71,177 | 11.57 |
| 78 | 0.047826 | 56,065 | 9.37 | 0.034321 | 68,981 | 10.92 |
| 79 | 0.052649 | 53,383 | 8.82 | 0.038211 | 66,613 | 10.29 |
| 80 | 0.058206 | 50,573 | 8.28 | 0.042771 | 64,068 | 9.68 |
| 81 | 0.064581 | 47,629 | 7.76 | 0.047992 | 61,328 | 9.09 |
| 82 | 0.071657 | 44,553 | 7.26 | 0.053678 | 58,385 | 8.52 |
| 83 | 0.079465 | 41,361 | 6.79 | 0.05981 | 55,251 | 7.98 |
| 84 | 0.088141 | 38,074 | 6.33 | 0.066584 | 51,946 | 7.45 |
| 85 | 0.097854 | 34,718 | 5.89 | 0.074258 | 48,487 | 6.95 |
| 86 | 0.108747 | 31,321 | 5.48 | 0.083053 | 44,887 | 6.47 |
| 87 | 0.120919 | 27,915 | 5.08 | 0.093123 | 41,159 | 6.01 |
| 88 | 0.134425 | 24,539 | 4.71 | 0.10454 | 37,326 | 5.57 |
| 89 | 0.149273 | 21,241 | 4.37 | 0.117305 | 33,424 | 5.16 |
| 90 | 0.165452 | 18,070 | 4.05 | 0.131392 | 29,503 | 4.78 |
| 91 | 0.182935 | 15,080 | 3.75 | 0.146753 | 25,627 | 4.43 |
| 92 | 0.201679 | 12,322 | 3.48 | 0.163331 | 21,866 | 4.11 |
| 93 | 0.221637 | 9,837 | 3.23 | 0.181064 | 18,294 | 3.81 |
| 94 | 0.242747 | 7,656 | 3.01 | 0.199886 | 14,982 | 3.55 |
| 95 | 0.263672 | 5,798 | 2.81 | 0.218908 | 11,987 | 3.31 |
| 96 | 0.284014 | 4,269 | 2.64 | 0.237815 | 9,363 | 3.09 |
| 97 | 0.303355 | 3,057 | 2.49 | 0.256265 | 7,136 | 2.9 |
| 98 | 0.321268 | 2,129 | 2.36 | 0.273894 | 5,308 | 2.73 |
| 99 | 0.337332 | 1,445 | 2.24 | 0.290328 | 3,854 | 2.58 |
| 100 | 0.354198 | 958 | 2.12 | 0.307747 | 2,735 | 2.42 |
| 101 | 0.371908 | 619 | 2.01 | 0.326212 | 1,893 | 2.28 |
| 102 | 0.390503 | 388 | 1.9 | 0.345785 | 1,276 | 2.14 |
| 103 | 0.410029 | 237 | 1.8 | 0.366532 | 835 | 2.01 |
| 104 | 0.43053 | 140 | 1.7 | 0.388524 | 529 | 1.88 |
| 105 | 0.452057 | 80 | 1.6 | 0.411835 | 323 | 1.76 |
| 106 | 0.474659 | 44 | 1.51 | 0.436546 | 190 | 1.65 |
| 107 | 0.498392 | 23 | 1.42 | 0.462738 | 107 | 1.54 |
| 108 | 0.523312 | 11 | 1.34 | 0.490503 | 58 | 1.44 |
| 109 | 0.549478 | 5 | 1.26 | 0.519933 | 29 | 1.34 |
| 110 | 0.576951 | 2 | 1.18 | 0.551129 | 14 | 1.24 |
| 111 | 0.605799 | 1 | 1.1 | 0.584196 | 6 | 1.15 |
| 112 | 0.636089 | 0 | 1.03 | 0.619248 | 3 | 1.06 |

| | | | | | | |
|---|---|---|---|---|---|---|
| 113 | 0.667893 | 0 | 0.96 | 0.656403 | 1 | 0.98 |
| 114 | 0.701288 | 0 | 0.9 | 0.695787 | 0 | 0.91 |
| 115 | 0.736353 | 0 | 0.84 | 0.736353 | 0 | 0.84 |
| 116 | 0.77317 | 0 | 0.78 | 0.77317 | 0 | 0.78 |
| 117 | 0.811829 | 0 | 0.72 | 0.811829 | 0 | 0.72 |
| 118 | 0.85242 | 0 | 0.66 | 0.85242 | 0 | 0.66 |
| 119 | 0.895041 | 0 | 0.61 | 0.895041 | 0 | 0.61 |

[a] Probability of dying within one year.

[b] Number of survivors out of 100,000 born alive.

Note: The period life expectancy at a given age for 2017 represents the average number of years of life remaining if a group of persons at that age were to experience the mortality rates for 2017 over the course of their remaining life.
