## Supplementary material for "2.5 Million Person-Years of Life Have Been Lost Due to COVID-19 in the United States": Table 11

**Table 11. Summary of current and projected person-years of life lost**

| Date | 10/3/20 | Projection 10/30/2020 (5) | Projection Est. 2/01/2021 (5) |
|---|---|---|---|
| Deaths with verified COVID-19 (1) | 194,087 | 230,000 | 400,000 |
| Estimated PYLL using average LE (2) | 2,572,102 | 3048032 | 5,300,925 |
| Estimated PYLL using adjusted LE (3) | 2,466,160 | 2922487 | 5,082,585 |
| Adjustment for co-morbidities, -15% (4) | 2,096,236 | 2,484,114 | 4,320,197 |

(1) Deaths for which a diagnosis of COVID-19 was obtained in medical records.

(2) Estimated using the LE (Life Expectancy) averaged over a decade.

(3) Estimated using the LE Adjusted for distribution within a decade and the error from combining the 95 and greater with the 85-94 group, 2,486,160- 20,000.

(4) Estimated correction to consider comorbidities.

(5) Estimates for projected numbers as of the indicated date.
