## Supplementary material for "2.5 Million Person-Years of Life Have Been Lost Due to COVID-19 in the United States": Table 10

| | A | B | C | D | E | F | G | H | I | J | K | L |
|---|---|---|---|---|---|---|---|---|---|---|---|---|
| 1 | **Table 10 Comorbidity distributions by age groups. From https://www.cdc.gov/nchs/nvss/vsrr/covid_weekly/index.htm#Comorbidities** | | | | | | | | | | | |
| 2 | | | | | | | | | | | | |
| 3 | **Updated October 14, 2020** | | | | | | | | | | | |
| 4 | | **Number of Conditions** | | | | | | | | | | |
| 5 | | **Age Group** | | | | | | | | | | |
| 6 | **Conditions Contributing to Deaths where COVID-19 was listed on the c** | **ICD–10 code** | **All ages** | **0–24 years** | **25–34 years** | **35–44 years** | **45–54 years** | **55–64 years** | **65–74 years** | **75–84 years** | **85 years and over** | |
| 7 | Total COVID-19 deaths[2], as of 10/10/2020 | U071 | 201,141 | 444 | 1,569 | 4,087 | 10,736 | 25,710 | 43,480 | 53,251 | 61,859 | |
| 8 | **Respiratory diseases** | | - | - | - | - | - | - | - | - | - | |
| 9 | Influenza and pneumonia | J09–J18 | 86,982 | 152 | 718 | 1,847 | 5,144 | 12,382 | 20,095 | 23,197 | 23,445 | |
| 10 | Chronic lower respiratory diseases | J40–J47 | 17,486 | 30 | 74 | 172 | 490 | 1,865 | 4,252 | 5,550 | 5,052 | |
| 11 | Adult respiratory distress syndrome | J80 | 26,196 | 75 | 293 | 761 | 2,176 | 4,703 | 6,956 | 6,334 | 4,896 | |
| 12 | Respiratory failure | J96 | 70,092 | 136 | 510 | 1,326 | 3,919 | 9,454 | 16,489 | 19,372 | 18,884 | |
| 13 | Respiratory arrest | R09.2 | 4,105 | 9 | 30 | 81 | 202 | 461 | 843 | 1,073 | 1,406 | |
| 14 | Other diseases of the respiratory system | J00–J06, J20- | 7,516 | 25 | 65 | 162 | 405 | 1,005 | 1,651 | 2,009 | 2,194 | |
| 15 | **Circulatory diseases** | | - | - | - | - | - | - | - | - | - | |
| 16 | Hypertensive diseases | I10–I15 | 43,801 | 21 | 128 | 575 | 1,895 | 5,333 | 9,700 | 12,079 | 14,070 | |
| 17 | Ischemic heart disease | I20–I25 | 22,903 | 4 | 31 | 129 | 553 | 2,125 | 4,747 | 6,964 | 8,349 | |
| 18 | Cardiac arrest | I46 | 24,653 | 61 | 240 | 591 | 1,630 | 3,644 | 5,670 | 6,151 | 6,666 | |
| 19 | Cardiac arrhythmia | I44, I45, I47– | 12,845 | 11 | 31 | 81 | 309 | 962 | 2,350 | 3,835 | 5,266 | |
| 20 | Heart failure | I50 | 13,426 | 6 | 49 | 105 | 355 | 1,144 | 2,348 | 3,740 | 5,679 | |
| 21 | Cerebrovascular diseases | I60–I69 | 9,947 | 7 | 31 | 106 | 372 | 1,144 | 2,268 | 2,883 | 3,135 | |
| 22 | Other diseases of the circulatory system | I00–I09, I26– | 11,494 | 54 | 123 | 270 | 654 | 1,515 | 2,585 | 2,882 | 3,411 | |
| 23 | **Sepsis** | A40–A41 | 18,815 | 45 | 177 | 465 | 1,411 | 3,301 | 5,225 | 4,729 | 3,462 | |
| 24 | **Malignant neoplasms** | C00–C97 | 9,386 | 32 | 44 | 127 | 358 | 1,277 | 2,461 | 2,802 | 2,285 | |
| 25 | **Diabetes** | E10–E14 | 33,100 | 56 | 214 | 760 | 2,269 | 5,513 | 8,970 | 8,815 | 6,503 | |
| 26 | **Obesity** | E65–E68 | 7,572 | 103 | 358 | 807 | 1,323 | 1,913 | 1,847 | 957 | 264 | |
| 27 | **Alzheimer disease** | G30 | 7,308 | 0 | 0 | 0 | 2 | 62 | 485 | 2,157 | 4,602 | |
| 28 | **Vascular and unspecified dementia** | F01, F03 | 22,696 | 0 | 0 | 2 | 24 | 348 | 2,134 | 6,763 | 13,425 | |
| 29 | **Renal failure** | N17–N19 | 18,031 | 23 | 150 | 378 | 1,173 | 2,770 | 4,684 | 4,705 | 4,146 | |
| 30 | **Intentional and unintentional injury, poisoning and other adverse even** | S00–T98, V0 | 6,871 | 52 | 199 | 255 | 432 | 829 | 1,351 | 1,642 | 2,111 | |
| 31 | **All other conditions and causes (residual)** | A00–A39, A4 | 102,277 | 341 | 955 | 2,108 | 5,644 | 13,890 | 24,026 | 26,830 | 28,475 | |

Notes

NOTE: Number of conditions reported in this table are tabulated from deaths received and coded as of the date of analysis and do not represent all deaths that occurred in that period. Data for this table are derived from a cut of the NVSS database taken at a particular time, separate from other surveillance tables on this page which are tabulated on the date of update. As a result, the total number of COVID-19 deaths in this table may not match other surveillance tables on this page.

1 Conditions contributing to the death were identified using the International Classification of Diseases, Tenth Revision (ICD–10). Deaths involving more than one condition (e.g., deaths involving both diabetes and respiratory arrest) were counted in both totals. To avoid counting the same death multiple times, the numbers for different conditions should not be summated.

2 Deaths with confirmed or presumed COVID-19, coded to ICD–10 code U07.1

3 Excess Deaths See the NCHS Excess Deaths Data Visualization.

This data visualization presents data on weekly counts of all-cause mortality by jurisdiction of occurrence. Counts of deaths in the most recent weeks are compared with historical trends to determine whether the number of deaths in recent weeks is significantly higher than expected.

Comorbidity distributions by age groups. From https://www.cdc.gov/nchs/nvss/vsrr/covid_weekly/index.htm#Comorbidities
