## Supplementary material for "2.5 Million Person-Years of Life Have Been Lost Due to COVID-19 in the United States": Table 9

Table 9 Actuarial Table for Sex, and Race in the US. Compiled from data in The National Vital Statistics Reports Volume 68, Number 7 June 24, 2019 United States Life Tables, 2917 by Elizabeth Arias and Jiaquan Xu, Division of Vital Statistics

Table 9A

| | White Males | White Females | Black Males | Black Females | Hispanic Males | Hispanic Females | Total US Popul. | All Whites | All Blacks | All Hispanics | Ave of Black and Hispanic | |
|---|---|---|---|---|---|---|---|---|---|---|---|---|
| | Expectation of life at age *x* | Expectation of life at age *x* | Expectation of life at age *x* | Expectation of life at age *x* | Expectation of life at age *x* | Expectation of life at age *x* | Expectation of life at age *x* | Expectation of life at age *x* | Expectation of life at age *x* | Expectation of life at age *x* | ExpAverage of Black and Hispanic at age *x* | Difference between all Whites and the average of all Black and Hispanics |
| Age (years) | $e_x$ | $e_x$ | $e_x$ | $e_x$ | $e_x$ | $e_x$ | $e_x$ | $e_x$ | $e_x$ | $e_x$ | $e_x$ | years |
| 0–1 | 76.1 | 81.0 | 71.5 | 78.1 | 79.1 | 84.3 | 78.5 | 78.8 | 75.3 | 81.8 | 78.5616798 | 0.0 |
| 1–2 | 75.5 | 80.3 | 71.4 | 77.9 | 78.5 | 83.7 | 77.9 | 78.2 | 75.1 | 81.2 | 78.1808243 | -0.3 |
| 2–3 | 74.5 | 79.3 | 70.4 | 76.9 | 77.5 | 82.7 | 76.9 | 77.2 | 74.2 | 80.3 | 77.2160225 | -0.3 |
| 3–4 | 73.5 | 78.4 | 69.4 | 75.9 | 76.6 | 81.7 | 75.9 | 76.2 | 73.2 | 79.3 | 76.2374496 | -0.3 |
| 4–5 | 72.6 | 77.4 | 68.5 | 75.0 | 75.6 | 80.8 | 74.9 | 75.2 | 72.2 | 78.3 | 75.2540703 | -0.3 |
| 5–6 | 71.6 | 76.4 | 67.5 | 74.0 | 74.6 | 79.8 | 74.0 | 74.2 | 71.2 | 77.3 | 74.2668343 | -0.3 |
| 6–7 | 70.6 | 75.4 | 66.5 | 73.0 | 73.6 | 78.8 | 73.0 | 73.2 | 70.3 | 76.3 | 73.2787743 | -0.3 |
| 7–8 | 69.6 | 74.4 | 65.5 | 72.0 | 72.6 | 77.8 | 72.0 | 72.3 | 69.3 | 75.3 | 72.2894478 | -0.3 |
| 8–9 | 68.6 | 73.4 | 64.5 | 71.0 | 71.6 | 76.8 | 71.0 | 71.3 | 68.3 | 74.3 | 71.299057 | -0.3 |
| 9–10 | 67.6 | 72.4 | 63.5 | 70.0 | 70.6 | 75.8 | 70.0 | 70.3 | 67.3 | 73.3 | 70.3076057 | -0.3 |
| 10–11 | 66.6 | 71.4 | 62.5 | 69.0 | 69.6 | 74.8 | 69.0 | 69.3 | 66.3 | 72.3 | 69.3151627 | -0.3 |
| 11–12 | 65.6 | 70.4 | 61.5 | 68.0 | 68.6 | 73.8 | 68.0 | 68.3 | 65.3 | 71.3 | 68.3221436 | -0.3 |
| 12–13 | 64.6 | 69.4 | 60.6 | 67.0 | 67.6 | 72.8 | 67.0 | 67.3 | 64.3 | 70.3 | 67.329525 | -0.3 |
| 13–14 | 63.6 | 68.4 | 59.6 | 66.1 | 66.6 | 71.8 | 66.0 | 66.3 | 63.3 | 69.4 | 66.3389454 | -0.3 |
| 14–15 | 62.6 | 67.4 | 58.6 | 65.1 | 65.7 | 70.8 | 65.0 | 65.3 | 62.3 | 68.4 | 65.352375 | -0.3 |
| 15–16 | 61.7 | 66.4 | 57.6 | 64.1 | 64.7 | 69.8 | 64.0 | 64.3 | 61.4 | 67.4 | 64.3712425 | -0.3 |
| 16–17 | 60.7 | 65.5 | 56.7 | 63.1 | 63.7 | 68.9 | 63.1 | 63.3 | 60.4 | 66.4 | 63.3960018 | -0.3 |
| 17–18 | 59.7 | 64.5 | 55.7 | 62.1 | 62.7 | 67.9 | 62.1 | 62.4 | 59.4 | 65.4 | 62.4265156 | -0.3 |
| 18–19 | 58.8 | 63.5 | 54.8 | 61.1 | 61.8 | 66.9 | 61.1 | 61.4 | 58.5 | 64.4 | 61.4627438 | -0.3 |
| 19–20 | 57.8 | 62.5 | 53.9 | 60.2 | 60.8 | 65.9 | 60.1 | 60.4 | 57.5 | 63.5 | 60.5043621 | -0.4 |
| 20–21 | 56.9 | 61.5 | 53.0 | 59.2 | 59.9 | 64.9 | 59.2 | 59.5 | 56.6 | 62.5 | 59.55093 | -0.4 |
| 21–22 | 55.9 | 60.6 | 52.1 | 58.2 | 58.9 | 64.0 | 58.2 | 58.5 | 55.7 | 61.6 | 58.6022968 | -0.4 |
| 22–23 | 55.0 | 59.6 | 51.2 | 57.3 | 58.0 | 63.0 | 57.3 | 57.6 | 54.7 | 60.6 | 57.6581116 | -0.4 |
| 23–24 | 54.1 | 58.6 | 50.3 | 56.3 | 57.0 | 62.0 | 56.3 | 56.6 | 53.8 | 59.6 | 56.7173996 | -0.4 |
| 24–25 | 53.1 | 57.7 | 49.4 | 55.3 | 56.1 | 61.0 | 55.4 | 55.7 | 52.9 | 58.7 | 55.7789841 | -0.4 |
| 25–26 | 52.2 | 56.7 | 48.5 | 54.4 | 55.2 | 60.1 | 54.5 | 54.7 | 51.9 | 57.7 | 54.8419342 | -0.4 |
| 26–27 | 51.3 | 55.7 | 47.6 | 53.4 | 54.2 | 59.1 | 53.5 | 53.8 | 51.0 | 56.8 | 53.9059353 | -0.4 |
| 27–28 | 50.4 | 54.8 | 46.8 | 52.5 | 53.3 | 58.1 | 52.6 | 52.9 | 50.1 | 55.8 | 52.9708538 | -0.4 |
| 28–29 | 49.5 | 53.8 | 45.9 | 51.5 | 52.4 | 57.1 | 51.7 | 51.9 | 49.2 | 54.9 | 52.0363693 | -0.4 |
| 29–30 | 48.6 | 52.8 | 45.0 | 50.6 | 51.4 | 56.2 | 50.7 | 51.0 | 48.3 | 53.9 | 51.1022568 | -0.4 |
| 30–31 | 47.7 | 51.9 | 44.1 | 49.6 | 50.5 | 55.2 | 49.8 | 50.0 | 47.4 | 53.0 | 50.1684704 | -0.4 |
| 31–32 | 46.8 | 50.9 | 43.2 | 48.7 | 49.6 | 54.2 | 48.9 | 49.1 | 46.4 | 52.0 | 49.2349606 | -0.4 |
| 32–33 | 45.9 | 50.0 | 42.3 | 47.7 | 48.6 | 53.2 | 47.9 | 48.2 | 45.5 | 51.1 | 48.3018837 | -0.4 |
| 33–34 | 45.0 | 49.0 | 41.4 | 46.8 | 47.7 | 52.3 | 47.0 | 47.3 | 44.6 | 50.1 | 47.3697128 | -0.4 |
| 34–35 | 44.1 | 48.1 | 40.6 | 45.9 | 46.8 | 51.3 | 46.1 | 46.3 | 43.7 | 49.2 | 46.438961 | -0.3 |
| 35–36 | 43.2 | 47.2 | 39.7 | 44.9 | 45.8 | 50.3 | 45.2 | 45.4 | 42.8 | 48.2 | 45.5099125 | -0.3 |
| 36–37 | 42.2 | 46.2 | 38.8 | 44.0 | 44.9 | 49.4 | 44.2 | 44.5 | 41.9 | 47.3 | 44.5828228 | -0.3 |
| 37–38 | 41.3 | 45.3 | 37.9 | 43.1 | 44.0 | 48.4 | 43.3 | 43.5 | 41.0 | 46.3 | 43.657671 | -0.3 |
| 38–39 | 40.4 | 44.3 | 37.1 | 42.1 | 43.0 | 47.4 | 42.4 | 42.6 | 40.1 | 45.4 | 42.7341442 | -0.3 |
| 39–40 | 39.5 | 43.4 | 36.2 | 41.2 | 42.1 | 46.5 | 41.5 | 41.7 | 39.2 | 44.4 | 41.8118954 | -0.3 |
| 40–41 | 38.6 | 42.5 | 35.3 | 40.3 | 41.2 | 45.5 | 40.6 | 40.8 | 38.3 | 43.5 | 40.8909054 | -0.3 |
| 41–42 | 37.7 | 41.5 | 34.5 | 39.4 | 40.3 | 44.5 | 39.7 | 39.8 | 37.4 | 42.6 | 39.9715462 | -0.3 |
| 42–43 | 36.8 | 40.6 | 33.6 | 38.5 | 39.3 | 43.6 | 38.7 | 38.9 | 36.5 | 41.6 | 39.0545044 | -0.3 |
| 43–44 | 35.9 | 39.7 | 32.8 | 37.6 | 38.4 | 42.6 | 37.8 | 38.0 | 35.6 | 40.7 | 38.1404705 | -0.3 |
| 44–45 | 35.1 | 38.7 | 31.9 | 36.7 | 37.5 | 41.7 | 36.9 | 37.1 | 34.7 | 39.7 | 37.2301064 | -0.3 |
| 45–46 | 34.2 | 37.8 | 31.0 | 35.8 | 36.6 | 40.7 | 36.0 | 36.2 | 33.8 | 38.8 | 36.3239212 | -0.3 |
| 46–47 | 33.3 | 36.9 | 30.2 | 34.9 | 35.7 | 39.8 | 35.1 | 35.3 | 33.0 | 37.9 | 35.4221478 | -0.3 |
| 47–48 | 32.4 | 36.0 | 29.4 | 34.0 | 34.8 | 38.8 | 34.2 | 34.4 | 32.1 | 37.0 | 34.5252018 | -0.3 |
| 48–49 | 31.5 | 35.1 | 28.5 | 33.1 | 33.8 | 37.9 | 33.3 | 33.5 | 31.2 | 36.0 | 33.6339397 | -0.3 |
| 49–50 | 30.7 | 34.2 | 27.7 | 32.2 | 32.9 | 36.9 | 32.4 | 32.6 | 30.4 | 35.1 | 32.7494507 | -0.3 |
| 50–51 | 29.8 | 33.3 | 26.9 | 31.4 | 32.1 | 36.0 | 31.6 | 31.7 | 29.5 | 34.2 | 31.8726845 | -0.3 |
| 51–52 | 28.9 | 32.4 | 26.1 | 30.5 | 31.2 | 35.1 | 30.7 | 30.8 | 28.7 | 33.3 | 31.0038948 | -0.3 |
| 52–53 | 28.1 | 31.5 | 25.3 | 29.7 | 30.3 | 34.2 | 29.8 | 29.9 | 27.9 | 32.4 | 30.1432676 | -0.3 |
| 53–54 | 27.3 | 30.6 | 24.5 | 28.8 | 29.4 | 33.2 | 29.0 | 29.1 | 27.1 | 31.5 | 29.2914867 | -0.3 |
| 54–55 | 26.4 | 29.7 | 23.7 | 28.0 | 28.6 | 32.3 | 28.1 | 28.2 | 26.2 | 30.7 | 28.4491653 | -0.3 |
| 55–56 | 25.6 | 28.8 | 23.0 | 27.2 | 27.7 | 31.4 | 27.3 | 27.4 | 25.5 | 29.8 | 27.616559 | -0.3 |
| 56–57 | 24.8 | 28.0 | 22.2 | 26.4 | 26.9 | 30.5 | 26.5 | 26.6 | 24.7 | 28.9 | 26.7933388 | -0.3 |
| 57–58 | 24.0 | 27.1 | 21.5 | 25.6 | 26.1 | 29.6 | 25.6 | 25.7 | 23.9 | 28.0 | 25.9792261 | -0.3 |
| 58–59 | 23.3 | 26.3 | 20.8 | 24.8 | 25.2 | 28.7 | 24.8 | 24.9 | 23.2 | 27.2 | 25.1745615 | -0.4 |

| | | | | | | | | | | | | | |
|---|---|---|---|---|---|---|---|---|---|---|---|---|---|
| 65 | 59-60 | 22.5 | 25.4 | 20.1 | 24.0 | 24.4 | 27.8 | 24.0 | 24.1 | 22.4 | 26.3 | 24.380084 | -0.4 |
| 66 | 60-61 | 21.7 | 24.6 | 19.4 | 23.3 | 23.6 | 27.0 | 23.2 | 23.3 | 21.7 | 25.5 | 23.5964432 | -0.4 |
| 67 | 61-62 | 21.0 | 23.8 | 18.7 | 22.5 | 22.8 | 26.1 | 22.4 | 22.5 | 21.0 | 24.7 | 22.8242331 | -0.4 |
| 68 | 62-63 | 20.2 | 22.9 | 18.1 | 21.7 | 22.0 | 25.2 | 21.6 | 21.7 | 20.3 | 23.9 | 22.0632095 | -0.4 |
| 69 | 63-64 | 19.5 | 22.1 | 17.4 | 21.0 | 21.3 | 24.4 | 20.9 | 20.9 | 19.6 | 23.0 | 21.3122721 | -0.5 |
| 70 | 64-65 | 18.7 | 21.3 | 16.8 | 20.3 | 20.5 | 23.5 | 20.1 | 20.2 | 18.9 | 22.2 | 20.5696335 | -0.5 |
| 71 | 65-66 | 18.0 | 20.5 | 16.2 | 19.5 | 19.7 | 22.7 | 19.3 | 19.4 | 18.2 | 21.4 | 19.8338652 | -0.5 |
| 72 | 66-67 | 17.3 | 19.7 | 15.6 | 18.8 | 19.0 | 21.9 | 18.6 | 18.6 | 17.6 | 20.6 | 19.10499 | -0.5 |
| 73 | 67-68 | 16.6 | 18.9 | 15.0 | 18.1 | 18.2 | 21.0 | 17.8 | 17.9 | 16.9 | 19.9 | 18.3835821 | -0.6 |
| 74 | 68-69 | 15.9 | 18.1 | 14.5 | 17.4 | 17.5 | 20.2 | 17.1 | 17.1 | 16.3 | 19.1 | 17.6692915 | -0.6 |
| 75 | 69-70 | 15.2 | 17.3 | 13.9 | 16.7 | 16.7 | 19.4 | 16.3 | 16.4 | 15.6 | 18.3 | 16.9614668 | -0.6 |
| 76 | 70-71 | 14.5 | 16.6 | 13.3 | 16.0 | 16.0 | 18.6 | 15.6 | 15.6 | 15.0 | 17.5 | 16.2603216 | -0.7 |
| 77 | 71-72 | 13.8 | 15.8 | 12.8 | 15.3 | 15.3 | 17.8 | 14.9 | 14.9 | 14.4 | 16.8 | 15.5650964 | -0.7 |
| 78 | 72-73 | 13.1 | 15.1 | 12.2 | 14.7 | 14.6 | 17.0 | 14.2 | 14.2 | 13.7 | 16.0 | 14.8796721 | -0.7 |
| 79 | 73-74 | 12.5 | 14.3 | 11.6 | 14.0 | 13.9 | 16.2 | 13.5 | 13.5 | 13.1 | 15.3 | 14.204493 | -0.7 |
| 80 | 74-75 | 11.8 | 13.6 | 11.1 | 13.4 | 13.2 | 15.4 | 12.8 | 12.9 | 12.5 | 14.6 | 13.5419073 | -0.7 |
| 81 | 75-76 | 11.2 | 12.9 | 10.6 | 12.7 | 12.6 | 14.7 | 12.2 | 12.2 | 11.9 | 13.8 | 12.8918419 | -0.7 |
| 82 | 76-77 | 10.6 | 12.2 | 10.1 | 12.1 | 11.9 | 13.9 | 11.5 | 11.5 | 11.4 | 13.1 | 12.254921 | -0.8 |
| 83 | 77-78 | 10.0 | 11.6 | 9.6 | 11.5 | 11.3 | 13.2 | 10.9 | 10.9 | 10.8 | 12.5 | 11.6316824 | -0.8 |
| 84 | 78-79 | 9.4 | 10.9 | 9.1 | 10.9 | 10.6 | 12.5 | 10.3 | 10.3 | 10.3 | 11.8 | 11.024394 | -0.8 |
| 85 | 79-80 | 8.8 | 10.3 | 8.6 | 10.3 | 10.0 | 11.8 | 9.7 | 9.7 | 9.7 | 11.1 | 10.4354234 | -0.8 |
| 86 | 80-81 | 8.3 | 9.7 | 8.1 | 9.8 | 9.4 | 11.1 | 9.1 | 9.1 | 9.2 | 10.5 | 9.8617506 | -0.8 |
| 87 | 81-82 | 7.8 | 9.1 | 7.7 | 9.3 | 8.8 | 10.4 | 8.5 | 8.5 | 8.7 | 9.9 | 9.30569887 | -0.8 |
| 88 | 82-83 | 7.3 | 8.5 | 7.3 | 8.7 | 8.3 | 9.8 | 8.0 | 8.0 | 8.2 | 9.3 | 8.76545429 | -0.8 |
| 89 | 83-84 | 6.8 | 7.9 | 6.9 | 8.2 | 7.7 | 9.2 | 7.5 | 7.5 | 7.8 | 8.7 | 8.24475932 | -0.8 |
| 90 | 84-85 | 6.3 | 7.4 | 6.5 | 7.7 | 7.2 | 8.6 | 7.0 | 7.0 | 7.3 | 8.2 | 7.74684405 | -0.8 |
| 91 | 85-86 | 5.9 | 6.9 | 6.1 | 7.3 | 6.7 | 8.0 | 6.5 | 6.5 | 6.9 | 7.6 | 7.26743531 | -0.8 |
| 92 | 86-87 | 5.4 | 6.4 | 5.7 | 6.8 | 6.3 | 7.5 | 6.0 | 6.1 | 6.5 | 7.1 | 6.80833721 | -0.8 |
| 93 | 87-88 | 5.0 | 5.9 | 5.4 | 6.4 | 5.8 | 6.9 | 5.6 | 5.6 | 6.1 | 6.6 | 6.36802578 | -0.8 |
| 94 | 88-89 | 4.7 | 5.5 | 5.1 | 6.0 | 5.4 | 6.4 | 5.2 | 5.2 | 5.7 | 6.2 | 5.95155072 | -0.8 |
| 95 | 89-90 | 4.3 | 5.1 | 4.8 | 5.6 | 5.0 | 6.0 | 4.8 | 4.8 | 5.4 | 5.7 | 5.55875015 | -0.7 |
| 96 | 90-91 | 4.0 | 4.7 | 4.5 | 5.2 | 4.6 | 5.5 | 4.5 | 4.5 | 5.1 | 5.3 | 5.18935442 | -0.7 |
| 97 | 91-92 | 3.7 | 4.3 | 4.2 | 4.9 | 4.3 | 5.1 | 4.1 | 4.1 | 4.8 | 4.9 | 4.84298491 | -0.7 |
| 98 | 92-93 | 3.4 | 4.0 | 3.9 | 4.6 | 4.0 | 4.7 | 3.8 | 3.8 | 4.5 | 4.6 | 4.51915956 | -0.7 |
| 99 | 93-94 | 3.1 | 3.7 | 3.7 | 4.3 | 3.7 | 4.4 | 3.5 | 3.5 | 4.2 | 4.2 | 4.21729779 | -0.7 |
| 100 | 94-95 | 2.9 | 3.4 | 3.5 | 4.0 | 3.4 | 4.0 | 3.3 | 3.3 | 3.9 | 3.9 | 3.9367274 | -0.7 |
| 101 | 95-96 | 2.7 | 3.2 | 3.3 | 3.7 | 3.2 | 3.7 | 3.0 | 3.0 | 3.7 | 3.6 | 3.67669392 | -0.6 |
| 102 | 96-97 | 2.5 | 2.9 | 3.1 | 3.5 | 2.9 | 3.4 | 2.8 | 2.8 | 3.5 | 3.4 | 3.43637192 | -0.6 |
| 103 | 97-98 | 2.3 | 2.7 | 2.9 | 3.3 | 2.7 | 3.2 | 2.6 | 2.6 | 3.3 | 3.2 | 3.21487546 | -0.6 |
| 104 | 98-99 | 2.2 | 2.5 | 2.8 | 3.0 | 2.5 | 3.0 | 2.4 | 2.4 | 3.1 | 2.9 | 3.01127172 | -0.6 |
| 105 | 99-100 | 2.0 | 2.3 | 2.6 | 2.9 | 2.4 | 2.7 | 2.3 | 2.3 | 2.9 | 2.7 | 2.82459283 | -0.5 |
| 106 | 100 and over | 1.9 | 2.2 | 2.5 | 2.7 | 2.2 | 2.6 | 2.1 | 2.1 | 2.7 | 2.6 | 2.65384984 | -0.5 |
| 107 | | | | | | | | | | | | Average Diff | -0.5 |

Vertical (Value) Axis Major Gridlines

**Table 9B**

| Row | Q | R | S | T | U | V | W |
|---|---|---|---|---|---|---|---|
| 3 | Males | White Males | Black Males | Hispanic Males | All US Males | | |
| 4 | | Expectation of life at age $x$ | Expectation of life at age $x$ | Expectation of life at age $x$ | Expectation of life at age $x$ | ExpAverage of Black and Hispanic at age $x$ | Difference between all Whites and the average of Male Black and Hispanics |
| 5 | Age (years) | $e_x$ | $e_x$ | $e_x$ | | | |
| 6 | 0–1 | 76.1 | 71.5 | 79.1 | 75.97 | **75.3033524** | **0.8** |
| 7 | 1–2 | 75.5 | 71.4 | 78.5 | 75.45 | **74.9462357** | **0.5** |
| 8 | 2–3 | 74.5 | 70.4 | 77.5 | 74.48 | **73.9807281** | **0.5** |
| 9 | 3–4 | 73.5 | 69.4 | 76.6 | 73.5 | **73.0024567** | **0.5** |
| 10 | 4–5 | 72.6 | 68.5 | 75.6 | 72.52 | **72.022007** | **0.5** |
| 11 | 5–6 | 71.6 | 67.5 | 74.6 | 71.53 | **71.0343742** | **0.5** |
| 12 | 6–7 | 70.6 | 66.5 | 73.6 | 70.54 | **70.0466576** | **0.5** |
| 13 | 7–8 | 69.6 | 65.5 | 72.6 | 69.55 | **69.0577774** | **0.5** |
| 14 | 8–9 | 68.6 | 64.5 | 71.6 | 68.56 | **68.0678139** | **0.5** |
| 15 | 9–10 | 67.6 | 63.5 | 70.6 | 67.57 | **67.0764847** | **0.5** |
| 16 | 10–11 | 66.6 | 62.5 | 69.6 | 66.57 | **66.083662** | **0.5** |
| 17 | 11–12 | 65.6 | 61.5 | 68.6 | 65.58 | **65.0898476** | **0.5** |
| 18 | 12–13 | 64.6 | 60.6 | 67.6 | 64.59 | **64.0966091** | **0.5** |
| 19 | 13–14 | 63.6 | 59.6 | 66.6 | 63.6 | **63.1066818** | **0.5** |
| 20 | 14–15 | 62.6 | 58.6 | 65.7 | 62.61 | **62.1233826** | **0.5** |
| 21 | 15–16 | 61.7 | 57.6 | 64.7 | 61.63 | **61.1490479** | **0.5** |
| 22 | 16–17 | 60.7 | 56.7 | 63.7 | 60.66 | **60.1842365** | **0.5** |
| 23 | 17–18 | 59.7 | 55.7 | 62.7 | 59.7 | **59.2285233** | **0.5** |
| 24 | 18–19 | 58.8 | 54.8 | 61.8 | 58.74 | **58.2817192** | **0.5** |
| 25 | 19–20 | 57.8 | 53.9 | 60.8 | 57.79 | **57.3431835** | **0.5** |
| 26 | 20–21 | 56.9 | 53.0 | 59.9 | 56.85 | **56.4121037** | **0.4** |
| 27 | 21–22 | 55.9 | 52.1 | 58.9 | 55.91 | **55.488184** | **0.4** |
| 28 | 22–23 | 55.0 | 51.2 | 58.0 | 54.98 | **54.5707493** | **0.4** |
| 29 | 23–24 | 54.1 | 50.3 | 57.0 | 54.06 | **53.6580658** | **0.4** |
| 30 | 24–25 | 53.1 | 49.4 | 56.1 | 53.14 | **52.7480698** | **0.4** |
| 31 | 25–26 | 52.2 | 48.5 | 55.2 | 52.22 | **51.8392086** | **0.4** |
| 32 | 26–27 | 51.3 | 47.6 | 54.2 | 51.31 | **50.9309711** | **0.4** |
| 33 | 27–28 | 50.4 | 46.8 | 53.3 | 50.39 | **50.0232143** | **0.4** |
| 34 | 28–29 | 49.5 | 45.9 | 52.4 | 49.48 | **49.1154232** | **0.4** |
| 35 | 29–30 | 48.6 | 45.0 | 51.4 | 48.56 | **48.2072277** | **0.4** |
| 36 | 30–31 | 47.7 | 44.1 | 50.5 | 47.65 | **47.2984982** | **0.4** |
| 37 | 31–32 | 46.8 | 43.2 | 49.6 | 46.74 | **46.3889942** | **0.4** |
| 38 | 32–33 | 45.9 | 42.3 | 48.6 | 45.83 | **45.478941** | **0.4** |
| 39 | 33–34 | 45.0 | 41.4 | 47.7 | 44.92 | **44.5693722** | **0.4** |
| 40 | 34–35 | 44.1 | 40.6 | 46.8 | 44.01 | **43.6615772** | **0.4** |
| 41 | 35–36 | 43.2 | 39.7 | 45.8 | 43.1 | **42.7563286** | **0.4** |
| 42 | 36–37 | 42.2 | 38.8 | 44.9 | 42.19 | **41.8540783** | **0.4** |
| 43 | 37–38 | 41.3 | 37.9 | 44.0 | 41.28 | **40.9545593** | **0.4** |
| 44 | 38–39 | 40.4 | 37.1 | 43.0 | 40.37 | **40.0569401** | **0.4** |
| 45 | 39–40 | 39.5 | 36.2 | 42.1 | 39.47 | **39.160162** | **0.4** |
| 46 | 40–41 | 38.6 | 35.3 | 41.2 | 38.56 | **38.2637157** | **0.4** |
| 47 | 41–42 | 37.7 | 34.5 | 40.3 | 37.65 | **37.3679161** | **0.4** |
| 48 | 42–43 | 36.8 | 33.6 | 39.3 | 36.75 | **36.4736767** | **0.4** |
| 49 | 43–44 | 35.9 | 32.8 | 38.4 | 35.85 | **35.5818443** | **0.4** |
| 50 | 44–45 | 35.1 | 31.9 | 37.5 | 34.95 | **34.6933012** | **0.4** |
| 51 | 45–46 | 34.2 | 31.0 | 36.6 | 34.06 | **33.8087416** | **0.4** |
| 52 | 46–47 | 33.3 | 30.2 | 35.7 | 33.17 | **32.9283676** | **0.4** |
| 53 | 47–48 | 32.4 | 29.4 | 34.8 | 32.28 | **32.0526838** | **0.3** |
| 54 | 48–49 | 31.5 | 28.5 | 33.8 | 31.41 | **31.1829138** | **0.3** |
| 55 | 49–50 | 30.7 | 27.7 | 32.9 | 30.54 | **30.3206491** | **0.3** |
| 56 | 50–51 | 29.8 | 26.9 | 32.1 | 29.67 | **29.4672432** | **0.3** |
| 57 | 51–52 | 28.9 | 26.1 | 31.2 | 28.82 | **28.6230936** | **0.3** |
| 58 | 52–53 | 28.1 | 25.3 | 30.3 | 27.98 | **27.7883797** | **0.3** |
| 59 | 53–54 | 27.3 | 24.5 | 29.4 | 27.14 | **26.9638653** | **0.3** |
| 60 | 54–55 | 26.4 | 23.7 | 28.6 | 26.32 | **26.1501818** | **0.3** |
| 61 | 55–56 | 25.6 | 23.0 | 27.7 | 25.5 | **25.34758** | **0.3** |
| 62 | 56–57 | 24.8 | 22.2 | 26.9 | 24.7 | **24.5556402** | **0.3** |
| 63 | 57–58 | 24.0 | 21.5 | 26.1 | 23.9 | **23.7741203** | **0.3** |
| 64 | 58–59 | 23.3 | 20.8 | 25.2 | 23.12 | **23.0036469** | **0.3** |

| | | | | | | | | |
|---|---|---|---|---|---|---|---|---|
| 65 | | 59-60 | 22.5 | 20.1 | 24.4 | 22.34 | **22.2454281** | **0.2** |
| 66 | | 60-61 | 21.7 | 19.4 | 23.6 | 21.58 | **21.5005388** | **0.2** |
| 67 | | 61-62 | 21.0 | 18.7 | 22.8 | 20.83 | **20.7698851** | **0.2** |
| 68 | | 62-63 | 20.2 | 18.1 | 22.0 | 20.08 | **20.0532484** | **0.2** |
| 69 | | 63-64 | 19.5 | 17.4 | 21.3 | 19.35 | **19.349349** | **0.1** |
| 70 | | 64-65 | 18.7 | 16.8 | 20.5 | 18.62 | **18.6559143** | **0.1** |
| 71 | | 65-66 | 18.0 | 16.2 | 19.7 | 17.89 | **17.9710398** | **0.0** |
| 72 | | 66-67 | 17.3 | 15.6 | 19.0 | 17.18 | **17.294744** | **0.0** |
| 73 | | 67-68 | 16.6 | 15.0 | 18.2 | 16.47 | **16.6274753** | **-0.1** |
| 74 | | 68-69 | 15.9 | 14.5 | 17.5 | 15.77 | **15.968348** | **-0.1** |
| 75 | | 69-70 | 15.2 | 13.9 | 16.7 | 15.07 | **15.3160167** | **-0.2** |
| 76 | | 70-71 | 14.5 | 13.3 | 16.0 | 14.39 | **14.6699071** | **-0.2** |
| 77 | | 71-72 | 13.8 | 12.8 | 15.3 | 13.71 | **14.0287099** | **-0.2** |
| 78 | | 72-73 | 13.1 | 12.2 | 14.6 | 13.05 | **13.3981166** | **-0.3** |
| 79 | | 73-74 | 12.5 | 11.6 | 13.9 | 12.4 | **12.77529** | **-0.3** |
| 80 | | 74-75 | 11.8 | 11.1 | 13.2 | 11.76 | **12.165278** | **-0.3** |
| 81 | | 75-76 | 11.2 | 10.6 | 12.6 | 11.14 | **11.5664611** | **-0.4** |
| 82 | | 76-77 | 10.6 | 10.1 | 11.9 | 10.53 | **10.9788218** | **-0.4** |
| 83 | | 77-78 | 10.0 | 9.6 | 11.3 | 9.94 | **10.405757** | **-0.4** |
| 84 | | 78-79 | 9.4 | 9.1 | 10.6 | 9.37 | **9.8447361** | **-0.4** |
| 85 | | 79-80 | 8.8 | 8.6 | 10.0 | 8.82 | **9.3024931** | **-0.5** |
| 86 | | 80-81 | 8.3 | 8.1 | 9.4 | 8.28 | **8.77698517** | **-0.5** |
| 87 | | 81-82 | 7.8 | 7.7 | 8.8 | 7.76 | **8.26571274** | **-0.5** |
| 88 | | 82-83 | 7.3 | 7.3 | 8.3 | 7.26 | **7.77055383** | **-0.5** |
| 89 | | 83-84 | 6.8 | 6.9 | 7.7 | 6.79 | **7.29851675** | **-0.5** |
| 90 | | 84-85 | 6.3 | 6.5 | 7.2 | 6.33 | **6.85039687** | **-0.5** |
| 91 | | 85-86 | 5.9 | 6.1 | 6.7 | 5.89 | **6.41752315** | **-0.6** |
| 92 | | 86-87 | 5.4 | 5.7 | 6.3 | 5.48 | **6.00002718** | **-0.6** |
| 93 | | 87-88 | 5.0 | 5.4 | 5.8 | 5.08 | **5.60069752** | **-0.6** |
| 94 | | 88-89 | 4.7 | 5.1 | 5.4 | 4.71 | **5.22507548** | **-0.6** |
| 95 | | 89-90 | 4.3 | 4.8 | 5.0 | 4.37 | **4.87281942** | **-0.6** |
| 96 | | 90-91 | 4.0 | 4.5 | 4.6 | 4.05 | **4.54347873** | **-0.6** |
| 97 | | 91-92 | 3.7 | 4.2 | 4.3 | 3.75 | **4.23649573** | **-0.6** |
| 98 | | 92-93 | 3.4 | 3.9 | 4.0 | 3.48 | **3.95121539** | **-0.6** |
| 99 | | 93-94 | 3.1 | 3.7 | 3.7 | 3.23 | **3.68689179** | **-0.5** |
| 100 | | 94-95 | 2.9 | 3.5 | 3.4 | 3.01 | **3.44270074** | **-0.5** |
| 101 | | 95-96 | 2.7 | 3.3 | 3.2 | 2.81 | **3.21775091** | **-0.5** |
| 102 | | 96-97 | 2.5 | 3.1 | 2.9 | 2.64 | **3.0110966** | **-0.5** |
| 103 | | 97-98 | 2.3 | 2.9 | 2.7 | 2.49 | **2.82175291** | **-0.5** |
| 104 | | 98-99 | 2.2 | 2.8 | 2.5 | 2.36 | **2.64870834** | **-0.5** |
| 105 | | 99-100 | 2.0 | 2.6 | 2.4 | 2.24 | **2.49093854** | **-0.4** |
| 106 | | 100 and over | 1.9 | 2.5 | 2.2 | 2.12 | **2.34741879** | **-0.4** |
| 107 | | | | | | | **Average Diff** | **0.1** |

| | X | Y | Z | AA | AB | AC | AD | AE | AF | AG | AH | AI |
|---|---|---|---|---|---|---|---|---|---|---|---|---|
| 1 | | | | | | | | | | | | |
| 2 | | | | | | | | | | | | |
| 3 | **Table 9C** | | **White Females** | **Black Females** | **Hispanic Females** | | | | | | | |
| 4 | | | Expectation of life at age $x$ | Expectation of life at age $x$ | Expectation of life at age $x$ | **ExpAverage of Black and Hispanic at age $x$** | **Difference between all Whites and the average of Female Black and Hispanics** | | | | | |
| 5 | | **Age (years)** | $e_x$ | $e_x$ | $e_x$ | | | | | | | |
| 6 | | 0–1 | 81.0 | 78.1 | 84.3 | **81.1932297** | **-0.2** | | | | | |
| 7 | | 1–2 | 80.3 | 77.9 | 83.7 | **80.7835197** | **-0.5** | | | | | |
| 8 | | 2–3 | 79.3 | 76.9 | 82.7 | **79.8201408** | **-0.5** | | | | | |
| 9 | | 3–4 | 78.4 | 75.9 | 81.7 | **78.8413544** | **-0.5** | | | | | |
| 10 | | 4–5 | 77.4 | 75.0 | 80.8 | **77.8550911** | **-0.5** | | | | | |
| 11 | | 5–6 | 76.4 | 74.0 | 79.8 | **76.8688545** | **-0.5** | | | | | |
| 12 | | 6–7 | 75.4 | 73.0 | 78.8 | **75.8806267** | **-0.5** | | | | | |
| 13 | | 7–8 | 74.4 | 72.0 | 77.8 | **74.8910294** | **-0.5** | | | | | |
| 14 | | 8–9 | 73.4 | 71.0 | 76.8 | **73.9003525** | **-0.5** | | | | | |
| 15 | | 9–10 | 72.4 | 70.0 | 75.8 | **72.908802** | **-0.5** | | | | | |
| 16 | | 10–11 | 71.4 | 69.0 | 74.8 | **71.9165764** | **-0.5** | | | | | |
| 17 | | 11–12 | 70.4 | 68.0 | 73.8 | **70.9239807** | **-0.5** | | | | | |
| 18 | | 12–13 | 69.4 | 67.0 | 72.8 | **69.9315262** | **-0.5** | | | | | |
| 19 | | 13–14 | 68.4 | 66.1 | 71.8 | **68.9399223** | **-0.5** | | | | | |
| 20 | | 14–15 | 67.4 | 65.1 | 70.8 | **67.9500046** | **-0.5** | | | | | |
| 21 | | 15–16 | 66.4 | 64.1 | 69.8 | **66.9623985** | **-0.5** | | | | | |
| 22 | | 16–17 | 65.5 | 63.1 | 68.9 | **65.9773922** | **-0.5** | | | | | |
| 23 | | 17–18 | 64.5 | 62.1 | 67.9 | **64.9950161** | **-0.5** | | | | | |
| 24 | | 18–19 | 63.5 | 61.1 | 66.9 | **64.0153179** | **-0.5** | | | | | |
| 25 | | 19–20 | 62.5 | 60.2 | 65.9 | **63.0381966** | **-0.5** | | | | | |
| 26 | | 20–21 | 61.5 | 59.2 | 64.9 | **62.0634613** | **-0.5** | | | | | |
| 27 | | 21–22 | 60.6 | 58.2 | 64.0 | **61.091135** | **-0.5** | | | | | |
| 28 | | 22–23 | 59.6 | 57.3 | 63.0 | **60.1211014** | **-0.5** | | | | | |
| 29 | | 23–24 | 58.6 | 56.3 | 62.0 | **59.1528854** | **-0.5** | | | | | |
| 30 | | 24–25 | 57.7 | 55.3 | 61.0 | **58.1858788** | **-0.5** | | | | | |
| 31 | | 25–26 | 56.7 | 54.4 | 60.1 | **57.2196655** | **-0.5** | | | | | |
| 32 | | 26–27 | 55.7 | 53.4 | 59.1 | **56.254076** | **-0.5** | | | | | |
| 33 | | 27–28 | 54.8 | 52.5 | 58.1 | **55.2891903** | **-0.5** | | | | | |
| 34 | | 28–29 | 53.8 | 51.5 | 57.1 | **54.3252087** | **-0.5** | | | | | |
| 35 | | 29–30 | 52.8 | 50.6 | 56.2 | **53.3625107** | **-0.5** | | | | | |
| 36 | | 30–31 | 51.9 | 49.6 | 55.2 | **52.401474** | **-0.5** | | | | | |
| 37 | | 31–32 | 50.9 | 48.7 | 54.2 | **51.4423866** | **-0.5** | | | | | |
| 38 | | 32–33 | 50.0 | 47.7 | 53.2 | **50.4853725** | **-0.5** | | | | | |
| 39 | | 33–34 | 49.0 | 46.8 | 52.3 | **49.5304489** | **-0.5** | | | | | |
| 40 | | 34–35 | 48.1 | 45.9 | 51.3 | **48.5774136** | **-0.5** | | | | | |
| 41 | | 35–36 | 47.2 | 44.9 | 50.3 | **47.6260567** | **-0.5** | | | | | |
| 42 | | 36–37 | 46.2 | 44.0 | 49.4 | **46.6764412** | **-0.5** | | | | | |
| 43 | | 37–38 | 45.3 | 43.1 | 48.4 | **45.7287636** | **-0.5** | | | | | |
| 44 | | 38–39 | 44.3 | 42.1 | 47.4 | **44.7830524** | **-0.5** | | | | | |
| 45 | | 39–40 | 43.4 | 41.2 | 46.5 | **43.8394432** | **-0.5** | | | | | |
| 46 | | 40–41 | 42.5 | 40.3 | 45.5 | **42.898243** | **-0.4** | | | | | |
| 47 | | 41–42 | 41.5 | 39.4 | 44.5 | **41.9598598** | **-0.4** | | | | | |
| 48 | | 42–43 | 40.6 | 38.5 | 43.6 | **41.0247116** | **-0.4** | | | | | |
| 49 | | 43–44 | 39.7 | 37.6 | 42.6 | **40.0931377** | **-0.4** | | | | | |
| 50 | | 44–45 | 38.7 | 36.7 | 41.7 | **39.1653137** | **-0.4** | | | | | |
| 51 | | 45–46 | 37.8 | 35.8 | 40.7 | **38.2413902** | **-0.4** | | | | | |
| 52 | | 46–47 | 36.9 | 34.9 | 39.8 | **37.3215179** | **-0.4** | | | | | |
| 53 | | 47–48 | 36.0 | 34.0 | 38.8 | **36.4060764** | **-0.4** | | | | | |
| 54 | | 48–49 | 35.1 | 33.1 | 37.9 | **35.4956532** | **-0.4** | | | | | |
| 55 | | 49–50 | 34.2 | 32.2 | 36.9 | **34.5910053** | **-0.4** | | | | | |
| 56 | | 50–51 | 33.3 | 31.4 | 36.0 | **33.6927776** | **-0.4** | | | | | |
| 57 | | 51–52 | 32.4 | 30.5 | 35.1 | **32.8010883** | **-0.4** | | | | | |
| 58 | | 52–53 | 31.5 | 29.7 | 34.2 | **31.9160624** | **-0.4** | | | | | |
| 59 | | 53–54 | 30.6 | 28.8 | 33.2 | **31.0382795** | **-0.5** | | | | | |
| 60 | | 54–55 | 29.7 | 28.0 | 32.3 | **30.1682825** | **-0.5** | | | | | |
| 61 | | 55–56 | 28.8 | 27.2 | 31.4 | **29.3062868** | **-0.5** | | | | | |
| 62 | | 56–57 | 28.0 | 26.4 | 30.5 | **28.4520416** | **-0.5** | | | | | |
| 63 | | 57–58 | 27.1 | 25.6 | 29.6 | **27.6052637** | **-0.5** | | | | | |
| 64 | | 58–59 | 26.3 | 24.8 | 28.7 | **26.7660875** | **-0.5** | | | | | |

| | | | | | | | |
|---|---|---|---|---|---|---|---|
| 65 | | 59–60 | 25.4 | 24.0 | 27.8 | **25.9349279** | **-0.5** |
| 66 | | 60–61 | 24.6 | 23.3 | 27.0 | **25.1121817** | **-0.5** |
| 67 | | 61–62 | 23.8 | 22.5 | 26.1 | **24.2982178** | **-0.5** |
| 68 | | 62–63 | 22.9 | 21.7 | 25.2 | **23.4928951** | **-0.6** |
| 69 | | 63–64 | 22.1 | 21.0 | 24.4 | **22.6955938** | **-0.6** |
| 70 | | 64–65 | 21.3 | 20.3 | 23.5 | **21.905324** | **-0.6** |
| 71 | | 65–66 | 20.5 | 19.5 | 22.7 | **21.1213379** | **-0.6** |
| 72 | | 66–67 | 19.7 | 18.8 | 21.9 | **20.3438883** | **-0.7** |
| 73 | | 67–68 | 18.9 | 18.1 | 21.0 | **19.5732813** | **-0.7** |
| 74 | | 68–69 | 18.1 | 17.4 | 20.2 | **18.8093767** | **-0.7** |
| 75 | | 69–70 | 17.3 | 16.7 | 19.4 | **18.0517941** | **-0.7** |
| 76 | | 70–71 | 16.6 | 16.0 | 18.6 | **17.3010912** | **-0.7** |
| 77 | | 71–72 | 15.8 | 15.3 | 17.8 | **16.557023** | **-0.8** |
| 78 | | 72–73 | 15.1 | 14.7 | 17.0 | **15.8218102** | **-0.8** |
| 79 | | 73–74 | 14.3 | 14.0 | 16.2 | **15.0991082** | **-0.8** |
| 80 | | 74–75 | 13.6 | 13.4 | 15.4 | **14.3886771** | **-0.8** |
| 81 | | 75–76 | 12.9 | 12.7 | 14.7 | **13.6920004** | **-0.8** |
| 82 | | 76–77 | 12.2 | 12.1 | 13.9 | **13.0101209** | **-0.8** |
| 83 | | 77–78 | 11.6 | 11.5 | 13.2 | **12.3414354** | **-0.8** |
| 84 | | 78–79 | 10.9 | 10.9 | 12.5 | **11.6918154** | **-0.8** |
| 85 | | 79–80 | 10.3 | 10.3 | 11.8 | **11.0599723** | **-0.8** |
| 86 | | 80–81 | 9.7 | 9.8 | 11.1 | **10.4426618** | **-0.8** |
| 87 | | 81–82 | 9.1 | 9.3 | 10.4 | **9.84588766** | **-0.8** |
| 88 | | 82–83 | 8.5 | 8.7 | 9.8 | **9.26537228** | **-0.8** |
| 89 | | 83–84 | 7.9 | 8.2 | 9.2 | **8.70263052** | **-0.8** |
| 90 | | 84–85 | 7.4 | 7.7 | 8.6 | **8.1626482** | **-0.8** |
| 91 | | 85–86 | 6.9 | 7.3 | 8.0 | **7.64458585** | **-0.7** |
| 92 | | 86–87 | 6.4 | 6.8 | 7.5 | **7.14251137** | **-0.7** |
| 93 | | 87–88 | 5.9 | 6.4 | 6.9 | **6.66003013** | **-0.7** |
| 94 | | 88–89 | 5.5 | 6.0 | 6.4 | **6.20348096** | **-0.7** |
| 95 | | 89–90 | 5.1 | 5.6 | 6.0 | **5.77286267** | **-0.7** |
| 96 | | 90–91 | 4.7 | 5.2 | 5.5 | **5.36803865** | **-0.7** |
| 97 | | 91–92 | 4.3 | 4.9 | 5.1 | **4.98873591** | **-0.7** |
| 98 | | 92–93 | 4.0 | 4.6 | 4.7 | **4.63454247** | **-0.6** |
| 99 | | 93–94 | 3.7 | 4.3 | 4.4 | **4.30491805** | **-0.6** |
| 100 | | 94–95 | 3.4 | 4.0 | 4.0 | **3.99919426** | **-0.6** |
| 101 | | 95–96 | 3.2 | 3.7 | 3.7 | **3.71659005** | **-0.6** |
| 102 | | 96–97 | 2.9 | 3.5 | 3.4 | **3.45622134** | **-0.5** |
| 103 | | 97–98 | 2.7 | 3.3 | 3.2 | **3.21711588** | **-0.5** |
| 104 | | 98–99 | 2.5 | 3.0 | 3.0 | **2.9982295** | **-0.5** |
| 105 | | 99–100 | 2.3 | 2.9 | 2.7 | **2.79846251** | **-0.5** |
| 106 | | 100 and over | 2.2 | 2.7 | 2.6 | **2.61667764** | **-0.4** |
| 107 | | | | | | **Average Diff** | **-0.6** |
