## Supplementary material for "2.5 Million Person-Years of Life Have Been Lost Due to COVID-19 in the United States": Table 8

| | A | B | C | D | E | F | G | H | I | J | K | L |
|---|---|---|---|---|---|---|---|---|---|---|---|---|
| 1 | **Table 8. Distribution of COVID-19 associated deaths among different ethnicities.** | | | | | | | | | | | |
| 2 | | | | | | | | | | | | |
| 3 | **Age group** | **Total COVID-19 Deaths** | **Non-Hispanic White** | **Non-Hispanic Black** | **Hispanic or Latino** | **Non-Hispanic American Indian** | **Non-Hispanic Asian[3]** | **Non-Hispanic Native Hawai** | **Non-Hispanic More than One Race** | **Unknown[4]** | | |
| 4 | All Ages | 198,809 | 102,250 | 41,066 | 42,295 | 1,987 | 8,188 | 341 | 561 | 2,121 | | |
| 5 | Under 1 year | 22 | 5 | 5 | 9 | 0 | 1 | 1 | 0 | 1 | | |
| 6 | 1–4 years | 15 | 5 | 3 | 5 | 1 | 0 | 0 | 1 | 0 | | |
| 7 | 5–14 years | 35 | 7 | 10 | 15 | 1 | 2 | 0 | 0 | 0 | | |
| 8 | 15–24 years | 369 | 60 | 116 | 163 | 9 | 10 | 4 | 4 | 3 | | |
| 9 | 25–34 years | 1,541 | 253 | 441 | 668 | 82 | 58 | 19 | 9 | 11 | | |
| 10 | 35–44 years | 4,039 | 589 | 1,113 | 2,007 | 115 | 142 | 25 | 16 | 32 | | |
| 11 | 45–54 years | 10,627 | 2,217 | 2,889 | 4,638 | 245 | 431 | 53 | 38 | 116 | | |
| 12 | 55–64 years | 25,421 | 7,974 | 7,141 | 8,267 | 420 | 1,097 | 90 | 90 | 342 | | |
| 13 | 65–74 years | 42,950 | 18,508 | 10,999 | 10,229 | 492 | 1,927 | 86 | 132 | 577 | | |
| 14 | 75–84 years | 52,618 | 29,781 | 10,402 | 9,196 | 386 | 2,076 | 42 | 145 | 590 | | |
| 15 | 85 years and | 61,172 | 42,851 | 7,947 | 7,098 | 236 | 2,444 | 21 | 126 | 449 | | |
| 16 | | | | | | | | | | | | |
| 17 | | | | | | | | | | | | |
| 18 | | | | | | | | | Distribution of deaths among races (%) | | | |
| 19 | **Age group** | **Total COVID-19 Deaths** | **White Non-Hispanic** | **Black Non-Hispanicor African American** | | **Hispanic or Latino** | **Sum of others** | | **White Nonhispanic** | **Black** | **Hispanic** | **Other** |
| 20 | All Ages | 198,809 | 102,250 | 41,066 | | 42,295 | 13,198 | | 51.43% | 20.07% | 21.27% | 6.64% |
| 21 | Under 1 year | 22 | 5 | 5 | | 9 | 3 | | | | | |
| 22 | 1–4 years | 15 | 5 | 3 | | 5 | 2 | | | | | |
| 23 | 5–14 years | 35 | 7 | 10 | | 15 | 3 | | | | | |
| 24 | 15–24 years | 369 | 60 | 116 | | 163 | 30 | | | | | |
| 25 | 25–34 years | 1,541 | 253 | 441 | | 668 | 179 | | | | | |
| 26 | 35–44 years | 4,039 | 589 | 1,113 | | 2,007 | 330 | | | | | |
| 27 | 45–54 years | 10,627 | 2,217 | 2,889 | | 4,638 | 883 | | | | | |
| 28 | 55–64 years | 25,421 | 7,974 | 7,141 | | 8,267 | 2,039 | | | | | |
| 29 | 65–74 years | 42,950 | 18,508 | 10,999 | | 10,229 | 3,214 | | | | | |
| 30 | 75–84 years | 52,618 | 29,781 | 10,402 | | 9,196 | 3,239 | | | | | |
| 31 | 85 years and | 61,172 | 42,851 | 7,947 | | 7,098 | 3,276 | | | | | |
