## Supplementary material for "2.5 Million Person-Years of Life Have Been Lost Due to COVID-19 in the United States": Table 7

**Table 7. Population distribution in the US by sex according to the 2000 and 2010 census.**

(For information on confidentiality protection, nonsampling error, and definitions, see www.census.gov/prod/cen2010/doc/sf1.pdf)

Adapted from Age and Sex Composition: 2010 Issued May 2011 2010 Census Briefs by Lindsay M. Howden and Julie A. Meyer https://www.census.gov/topics/population/age-and-sex/library/publications.2011.html

**Population by Age and Sex: 2000 and 2010**

| Census Year | 2000 | 2000 | 2000 | 2010 | 2010 | 2010 | Percent change, 2000 to 2010 | Percent change, 2000 to 2010 | Percent change, 2000 to 2010 |
|---|---|---|---|---|---|---|---|---|---|
| | Both sexes | Male | Female | Both sexes | Male | Female | Both sexes | Male | Female |
| **All ages** | **281,421,906** | **138,053,563** | **143,368,343** | **308,745,538** | **151,781,326** | **156,964,212** | **9.7** | **9.9** | **9.5** |
| Under 5 years . | 19,175,798 | 9,810,733 | 9,365,065 | 20,201,362 | 10,319,427 | 9,881,935 | 5.3 | 5.2 | 5.5 |
| 5 to 9 years. | 20,549,505 | 10,523,277 | 10,026,228 | 20,348,657 | 10,389,638 | 9,959,019 | –1.0 | –1.3 | –0.7 |
| 10 to 14 years. | 20,528,072 | 10,520,197 | 10,007,875 | 20,677,194 | 10,579,862 | 10,097,332 | 0.7 | 0.6 | 0.9 |
| 15 to 19 years. | 20,219,890 | 10,391,004 | 9,828,886 | 22,040,343 | 11,303,666 | 10,736,677 | 9.0 | 8.8 | 9.2 |
| 20 to 24 years. | 18,964,001 | 9,687,814 | 9,276,187 | 21,585,999 | 11,014,176 | 10,571,823 | 13.8 | 13.7 | 14.0 |
| 25 to 29 years. | 19,381,336 | 9,798,760 | 9,582,576 | 21,101,849 | 10,635,591 | 10,466,258 | 8.9 | 8.5 | 9.2 |
| 30 to 34 years. | 20,510,388 | 10,321,769 | 10,188,619 | 19,962,099 | 9,996,500 | 9,965,599 | [illegible] | –3.2 | –2.2 |
| 35 to 39 years. | 22,706,664 | 11,318,696 | 11,387,968 | 20,179,642 | 10,042,022 | 10,137,620 | –11.1 | –11.3 | –11.0 |
| 40 to 44 years. | 22,441,863 | 11,129,102 | 11,312,761 | 20,890,964 | 10,393,977 | 10,496,987 | –6.9 | –6.6 | –7.2 |
| 45 to 49 years. | 20,092,404 | 9,889,506 | 10,202,898 | 22,708,591 | 11,209,085 | 11,499,506 | 13.0 | 13.3 | 12.7 |
| 50 to 54 years. | 17,585,548 | 8,607,724 | 8,977,824 | 22,298,125 | 10,933,274 | 11,364,851 | 26.8 | 27.0 | 26.6 |
| 55 to 59 years. | 13,469,237 | 6,508,729 | 6,960,508 | 19,664,805 | 9,523,648 | 10,141,157 | 46.0 | 46.3 | 45.7 |
| 60 to 64 years. | 10,805,447 | 5,136,627 | 5,668,820 | 16,817,924 | 8,077,500 | 8,740,424 | 55.6 | 57.3 | 54.2 |
| 65 to 69 years. | 9,533,545 | 4,400,362 | 5,133,183 | 12,435,263 | 5,852,547 | 6,582,716 | 30.4 | 33.0 | 28.2 |
| 70 to 74 years. | 8,857,441 | 3,902,912 | 4,954,529 | 9,278,166 | 4,243,972 | 5,034,194 | 4.7 | 8.7 | 1.6 |
| 75 to 79 years. | 7,415,813 | 3,044,456 | 4,371,357 | 7,317,795 | 3,182,388 | 4,135,407 | –1.3 | 4.5 | –5.4 |
| 80 to 84 years. | 4,945,367 | 1,834,897 | 3,110,470 | 5,743,327 | 2,294,374 | 3,448,953 | 16.1 | 25.0 | 10.9 |
| 85 to 89 years. | 2,789,818 | 876,501 | 1,913,317 | 3,620,459 | 1,273,867 | 2,346,592 | 29.8 | 45.3 | 22.6 |
| 90 to 94 years. | 1,112,531 | 282,325 | 830,206 | 1,448,366 | 424,387 | 1,023,979 | 30.2 | 50.3 | 23.3 |
| 95 to 99 years. | 286,784 | 58,115 | 228,669 | 371,244 | 82,263 | 288,981 | 29.5 | 41.6 | 26.4 |
| 100 years and over . | 50,454 | 10,057 | 40,397 | 53,364 | 9,162 | 44,202 | 5.8 | –8.9 | 9.4 |
| Median age. | 35.3 | 34.0 | 36.5 | 37.2 | 35.8 | 38.5 | (X) | (X) | (X) |
